# Genetic Architecture and Risk Prediction of Gestational Diabetes Mellitus in over 116,144 Chinese Pregnancies

**DOI:** 10.1101/2025.03.14.25323926

**Authors:** Yuqin Gu, Hao Zheng, Piao Wang, Yanhong Liu, Xinxin Guo, Yuandan Wei, Zijing Yang, Shiyao Cheng, Yanchao Chen, Liang Hu, Xiaohang Chen, Quanfu Zhang, Guobo Chen, Fengxiang Wei, Jianxin Zhen, Siyang Liu

## Abstract

Gestational diabetes mellitus (GDM), a heritable metabolic disorder and the most common pregnancy-related condition, remains understudied regarding its genetic architecture and its potential for early prediction using genetic data. Here we conducted genome-wide association studies on 116,144 Chinese pregnancies, leveraging their non-invasive prenatal test (NIPT) sequencing data and detailed prenatal records. We identified 13 novel loci for GDM and 111 for five glycemic traits, with minor allele frequencies of 0.01-0.5 and absolute effect sizes of 0.03-0.62. Approximately 50% of these loci were specific to GDM and gestational glycemic levels, distinct from type 2 diabetes and general glycemic levels in East Asians. A machine learning model integrating polygenic risk scores (PRS) and prenatal records predicted GDM before 20 weeks of gestation, achieving an AUC of 0.729 and an accuracy of 0.835. Shapley values highlighted PRS as key contributors. This model offers a cost-effective strategy for early GDM prediction using clinical NIPT.

## Introduction

Gestational diabetes mellitus (GDM) is the most prevalent metabolic disorder in pregnancy, affecting 14% of pregnancies globally^1^. Elevated blood glucose levels in GDM substantially increase the risk of short-and long-term complications for both mother and offspring^1, 2, 3^. Short-term complications include neonatal hypoglycemia, preeclampsia, and birth trauma^4^, while long-term risks include a strong association with type 2 diabetes (T2D), affecting 2.5%-16.7% of women within one year postpartum and over 40% within ten years^5^. Recent randomized controlled trials (RCTs) have demonstrated that early prediction and prevention of GDM, particularly through interventions initiated in the first or early second trimesters, can significantly reduce adverse pregnancy outcomes^6^. However, effective and cost-efficient methods for early GDM detection remain scarce.

Despite the high heritability of GDM and the potential values of genetic variation in understanding its pathogenesis^7^, knowledge of GDM genetics remains limited, particularly in under-represented East Asian populations. Genome-wide association studies (GWAS) on GDM in East Asians are rare, with the most comprehensive study to date being our previous research, which identified four loci associated with GDM in 3,317 cases and 19,565 controls^8^. Limited understanding of the genetic architecture of GDM and related phenotypes hinders the identification of key genes and molecular pathways, thereby impeding advances in precise diagnosis and treatment^9^. Furthermore, existing GDM prediction models primarily rely on retrospective electronic health records data^10^ or a few SNPs identified in T2D GWAS^11^. These models do not incorporate polygenic risk information, despite the polygenic nature of GDM.

In this study, we analyzed large-scale sequencing data from non-invasive prenatal tests (NIPT) from 116,144 pregnant participants using methodologies we developed^12, 13^. We incorporated NIPT data with glycemic traits and comprehensive laboratory and electronic medical records (**Table S1**) to investigate the genetic architecture of GDM and the related glycemic traits. Specifically, we conducted a GWAS on 12,024 GDM cases (15.2%) and 67,845 non-diabetes controls (84.8%), as well as five glycemic traits: fasting plasma glucose (FPG, *n* = 56, 912), oral glucose tolerance test at 0 hours (OGTT0H, *n* = 85,086), 1 hour (OGTT1H, *n* = 85,956), 2 hours (OGTT2H, *n* = 84,743) and hemoglobin A1c (HbA1c, n = 69,269) (**Figure 1 and Figure 2**). The GWAS results were replicated in two independent cohorts: the Baoan 20K cohort (n = 20,439) and the NIPT PLUS cohort (n = 5,897) (**Supplementary Notes**).

**Figure 1.**
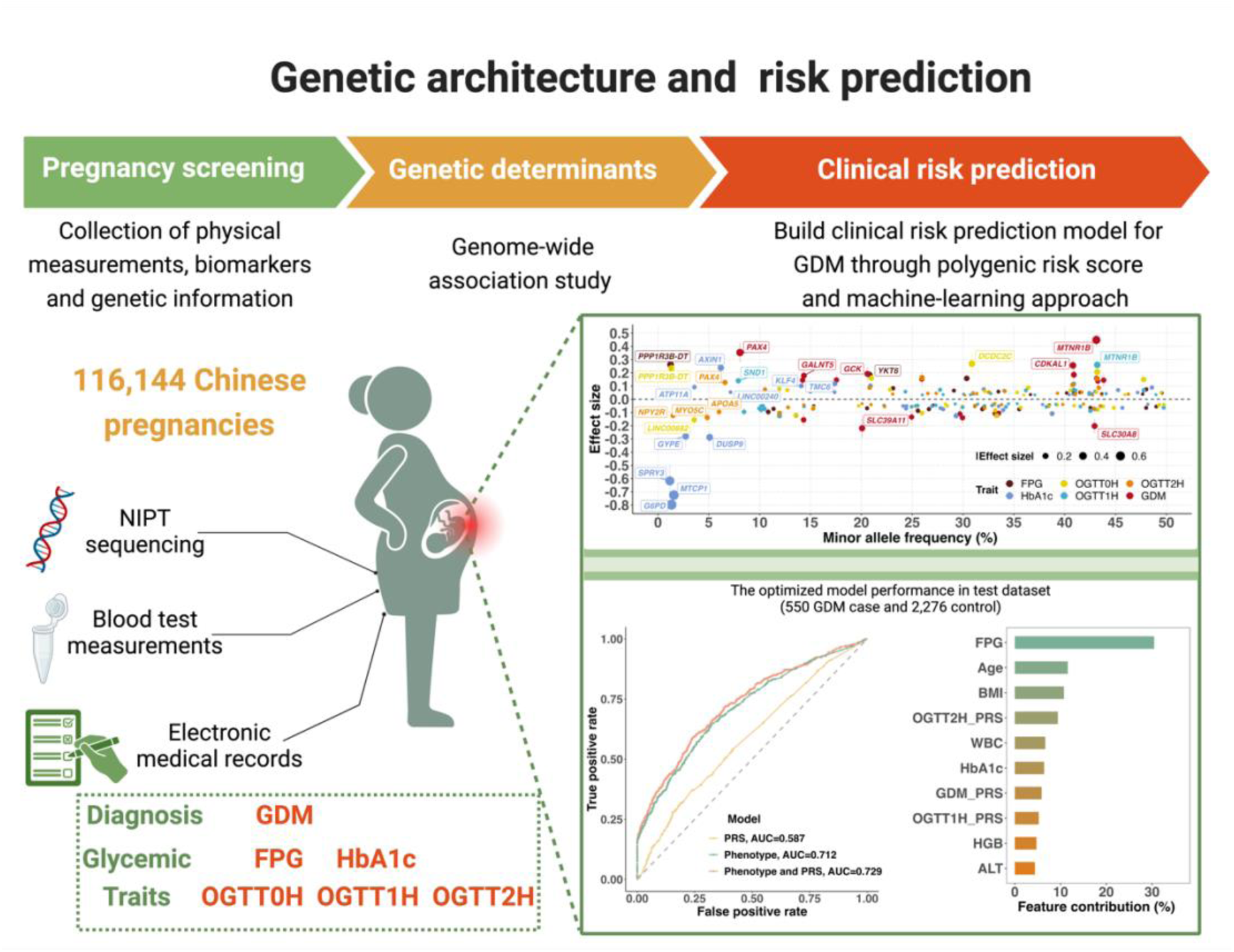
Study design, depicting the data sources and analyses performed. GDM: gestational diabetes mellitus; FPG: fasting plasma glucose; OGTT0H, 1H and 2H: oral glucose tolerance test at 0, 1 hour and 2 hours; WBC: white blood cell; HbA1c: glycated hemoglobin; HGB: hemoglobin concentration; ALT: alanine transaminase; PRS: polygenetic risk score.

**Figure 2.**
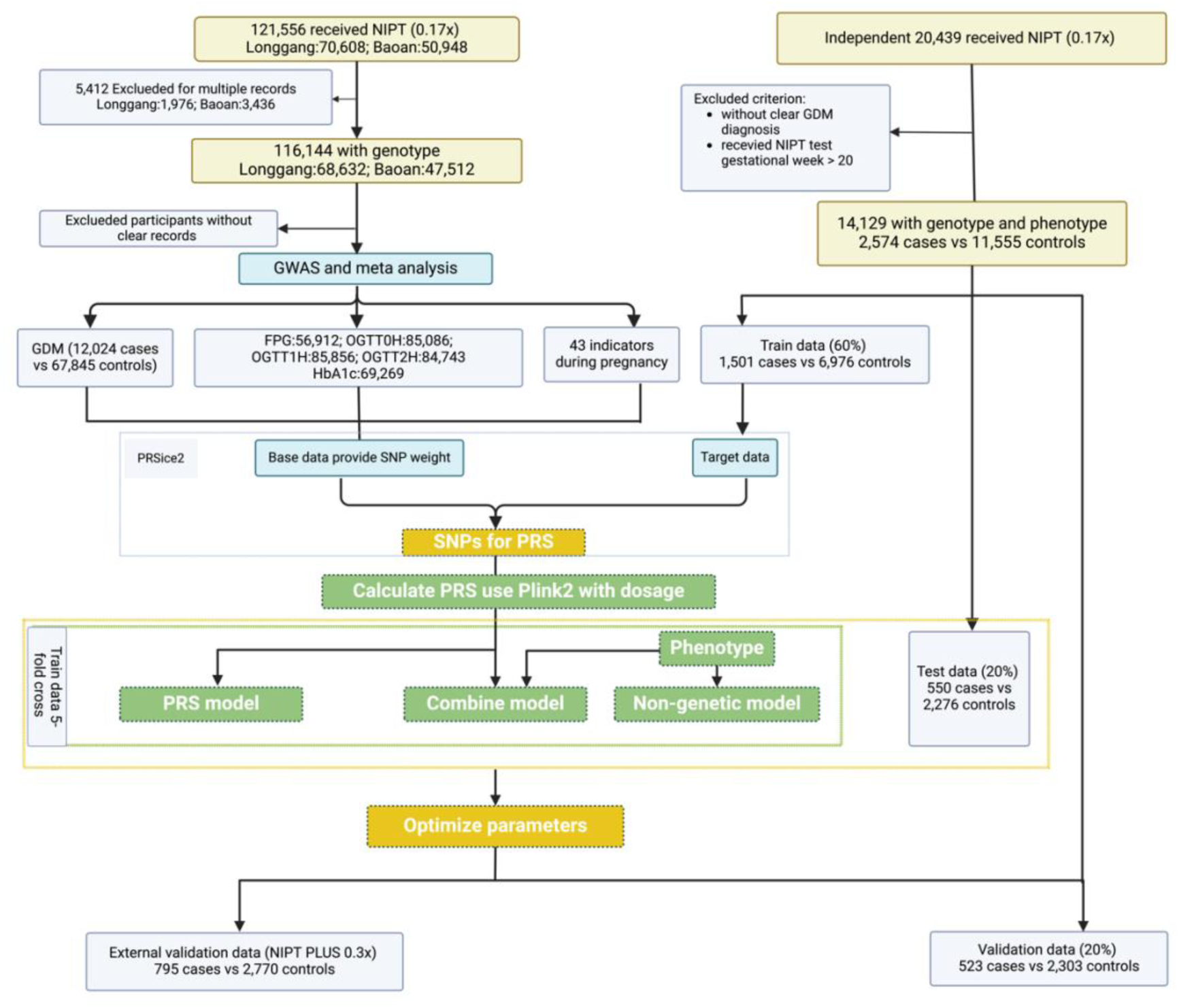
Flow diagram of participant quality control, GWAS, and the GDM early-prediction model. GDM: gestational diabetes mellitus; FPG: fasting plasma glucose; OGTT0H, 1H and 2H: oral glucose tolerance test at 0, 1 hour and 2 hours; HbA1c: glycated hemoglobin; SNP: single nucleotide polymorphism; PRS: polygenetic risk score.

Using these large-scale GWAS results, we developed and validated a machine learning model that integrates polygenic risk scores (PRS) with GDM and common biomarkers, as well as electronic health records from early pregnancy to predict GDM risk before 20 weeks of gestation (**Figure 1 and Figure 2**). This model utilized existing NIPT data and early prenatal screening information without incurring additional experimental costs, aside from minimal computational expenses. Our findings elucidate the genetic architecture of GDM and related glycemic traits in East Asians and provide a cost-effective model for early GDM prediction leveraging routine clinical NIPT data.

## Results

### Large-scale genome-wide association study of GDM

We conducted a GWAS on 116,144 pregnant women, including 12, 024 cases and 67,845 controls (**Figure 2**), using three analytical methods: PLINK2, REGENIE and BOLT-LMM (**Methods**). The results indicate that genetic effect estimates for the lead SNPs are highly consistent across methods (**Figure S1-2, Table S2**). Among the three approaches, PLINK2 yielded the expected LDSC intercepts and ratios for the highest proportion of phenotypes (**Table S3**). Therefore, we selected PLINK2 as the primary method for reporting results. We identified 19 independent loci significantly associated with GDM (**Figure 3A**). This represents a nearly fivefold increase compared to loci previously identified in East Asian populations^8, 14, 15, 16, 17^. Negligible statistical inflation was observed (QQ plot and LDSC intercept, **Figure S3)**. Excluding BMI as a covariate yielded consistent results **(Figure S4A, Table S4)**. The genetic architecture of GDM is highly polygenic, with all lead SNPs being common (MAF > 0.08) and absolute effect sizes ranging from 0.11 to 0.45. The *MTNR1B* locus displayed the strongest genetic effect (OR = 1.57, 95% CI [1.53, 1.60]). Notably, thirteen loci were identified for the first time, even when compared to recent European GDM GWAS findings^18^ **(highlighted in red, Figure 3A; Table S4-5)**. Power analysis is presented in **Figure S5**. SNP heritability (ℎ^2^) for GDM was estimated at 6.9% (s.e. 0.7%) (**Table S6**). Strong genetic correlations were observed between GDM and East Asian female T2D^19^ (*r_g_* = 0.525), and European GDM^18^ (*r_g_* = 0.439). Regional association plots showing all genes in a 1-Mbp window of the lead SNP are presented in **Figure S6**.

**Figure 3.**
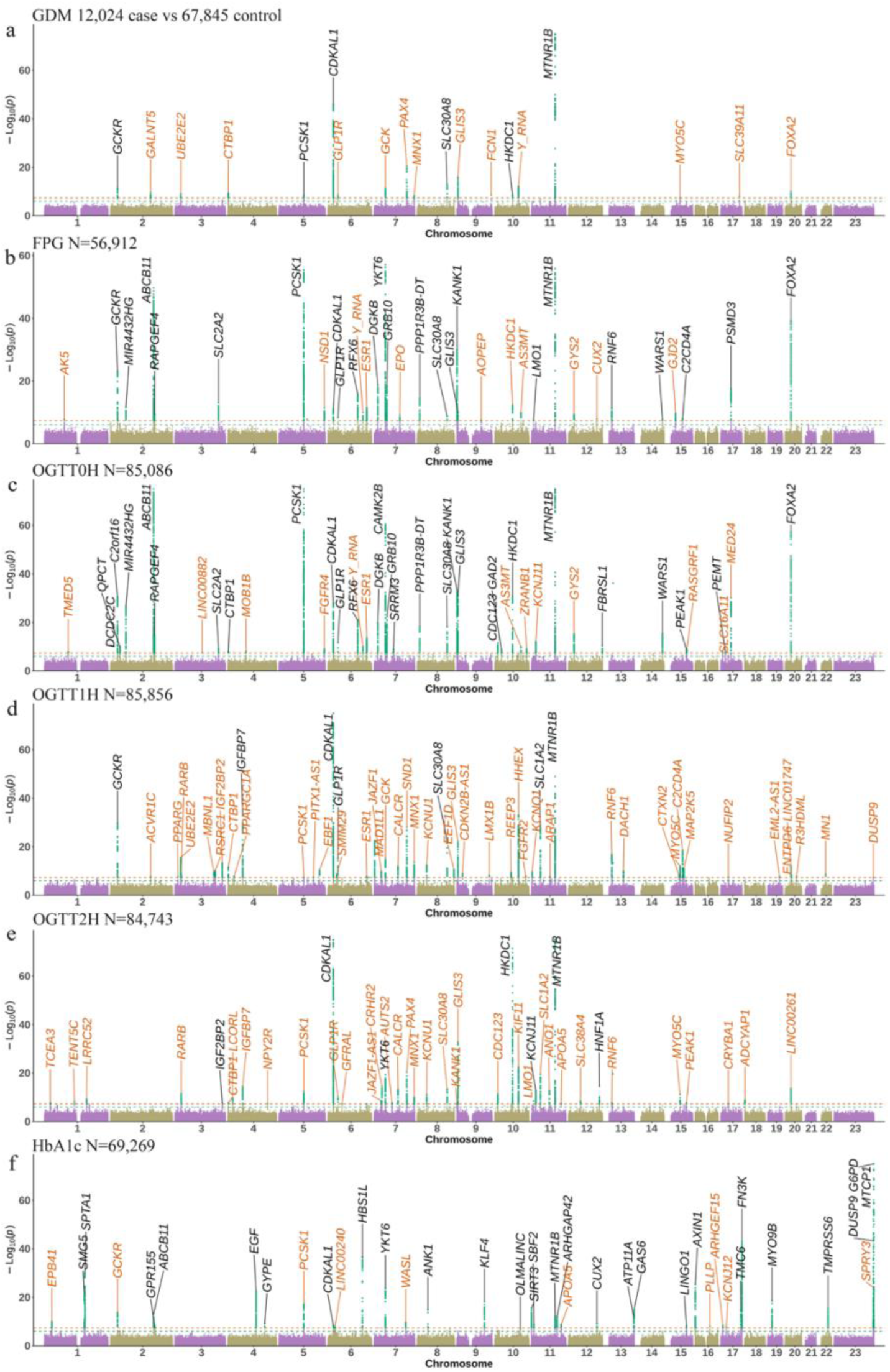
Manhattan and QQ plots for GDM and the five glycemic traits during gestation. Panels A-F display the GWAS results for GDM, FPG, OGTT0H, OGTT1H, OGTT2H and HbA1c, respectively. Horizontal lines indicate genome-wide significance (*P* = 5 ×10^−8^; red) and suggestive significance (*P* = 1 × 10^−6^; green) thresholds. Black: previously reported loci in GWAS Catalog (accessed on 2025/2/18); red: newly identified loci.

To validate these findings, we compared effect estimates between the two hospitals and with two replication cohorts (Baoan 20K and NIPT PLUS; details in **Supplementary Notes**). All loci exhibited consistent effect directions in the Longgang and Baoan cohorts, with 17 of 19 loci having p-values < 0.05 (**Figure S7A, Table S7**). In the meta-analysis of Baoan 20K and NIPT PLUS, 10 loci were replicated, and 18 demonstrated consistent effect directions (**Figure S8A, Table S8**). Furthermore, when compared to European GDM cohorts^18^, 10 loci were replicated, and 16 loci demonstrated consistent effect directions (**Figure S9A**, **Table S5**).

### Genome-wide association of five glycemic traits

Furthermore, we identified 205 independent loci significantly associated with five glycemic traits (*P* < 5×10^-8^): 34 for FPG, 42 for OGTT0H, 50 for OGTT1H, 41 for OGTT2H and 38 for HbA1c. Of these, 111 loci were newly discovered, with 11 associated with FPG,13 with OGTT0H, 43 with OGTT1H, 34 with OGTT2H and 10 with HbA1c (**Figure 3B-E, Table S4)**. The QQ plot and LDSC intercepts indicated negligible statistical inflation **(Figure S3)**, and results excluding BMI as covariates were also highly consistent (**Figure S4B, Table S4**). The genetic architecture of these traits was highly polygenic, with MAF ranging from 0.01 to 0.5 and absolute effect sizes between 0.03 and 0.73. Several loci had MAFs between 1% and 5%, demonstrating notable effect sizes (**Figure 1**). SNP heritability estimates for these traits ranged from 11.2% to 17.0% (s.e. ∼1.0%) (**Table S6**).

Of the identified loci, 197 (96.1%) exhibited consistent effect directions between the two hospitals (**Figure S7B-F, Table S7**), and 199 loci (97.1%) showed consistent effect directions compared to the meta-analysis of the Baoan 20K and NIPT PLUS cohorts, with 110 (53.7%) loci replicated using the Bonferroni criterion (**Figure S8B-F, Table S8**). Compared with the East Asian summary statistics from the Taiwan biobank, 25/34 loci for FPG and 25/38 loci for HbA1c had consistent effects and p-values < 0.05. Compared with East Asian summary statistics from the MAGIC consortium, 9/41 loci for OGTT2H showed consistent genetic effects and p-values < 0.05 (**Figure S9B-D, Table S5**). Upon further analysis, we found that the observed differences for OGTT2H were likely due to reduced statistical power in the MAGIC study (**Figure S10)**. Additional factors, including population stratification, phenotypic heterogeneity, and methodological differences between our study and the MAGIC consortium may also have contributed to these discrepancies **(Table S9)**.

Key biological findings include: (1) *MTNR1B* remained the most significant signal across all glycemic traits except HbA1c. For OGTT1H and OGTT2H, *MTNR1B* was not previously reported, apart from findings in our earlier study^8, 20^; (2) The genetic determinants of FPG (examined at ∼16 gestational week) and OGTT0H (measured between 24 and 28 gestational week) were highly similar, sharing 22 significant loci. The lead SNP effect sizes were strongly correlated (Pearson’s r = 0.975, *P* < 2.2⊆10^-16^), with a genetic correlation of 0.943(0.026), *P* = 9.33⊆10^-289^ (**Figure 3B-C, Figure S11-12, Table S10**); (3) Genetic determinants for baseline glycemic levels (FPG and OGTT0H) differed substantially from those after challenge (OGTT0H and OGTT2H). For example, *ABCB11* (lead SNP rs74870851), associated with severe cholestatic liver disease, and *FOXA2* (lead SNP rs6048206), linked to maturity-onset diabetes of the young, were specific to baseline glycemic traits but not OGTT1H and OGTT2H (**Figure 3C-E, Table S4**); (4) Several loci significantly associated with glycemic traits, including *ABCB11, YKT6* and *KANK1*, did not influence GDM susceptibility (**Figure 3**); (5) Genetic determinants of HbA1c, including *C2orf16*, *ABCA11, PCSK1, YKT6,* and *MTNR1B,* were also associated with glycemic traits. The most significant locus, *MTCP1*(lead SNP rs182573065) on the X chromosome **(Figure 3)**, was linked to venous thromboembolism, a condition related to blood cells such as platelets. Other loci were associated with various type of blood cells^21^, with the exceptions of *GYPE, OLMALINC* and *SIRT3*.

These findings highlight the polygenic nature of glycemic traits and their shared but distinct genetic determinants with related metabolic and hematological conditions.

### Distinct and shared variants between East Asian GDM and T2D

We explored the genetic relationship between Chinese GDM and East Asian Female T2D in more details using a Bayesian classification algorithm^22^ and the ‘coloc’ package (**Figure S13**). Our analysis identified two groups of GDM-associated loci: one shared with T2D and another predominantly associated with GDM (**Figure 4A, Table S11**). The GDM-predominant group comprised 12 of the 19 loci, which exhibited effect sizes approximately ten times greater in GDM compared to T2D. These loci also demonstrated a posterior probability of having the same single causal SNP (PPH4) below 0.5 in the *coloc* analysis (**Figure 4A, Table S11**). In contrast, the second group exhibited similar genetic effects between Chinese GDM and East Asian Female T2D, with PPH4 of at least 0.2 (**Figure 4A, Table S11**). Consistent with findings from European studies^18^, the *GCKR*, *PCSK1* and *MTNR1B* loci demonstrated a GDM-predominant effect. However, in contrast to European populations, where *CDKAL1* was identified as T2D-predominant^18^, this locus was shared between GDM and Female T2D in the East Asian population. Pathway enrichment analysis comparing the 12 GDM-predominant loci revealed associations with four categories of pathways: maturity onset diabetes of the young (*P* = 1.38 × 10^-4^), regulation of protein secretion (*P* = 1.75 × 10^-3^), glucose homeostasis (*P* = 2.78 × 10^-3^) and protein import into cell (*P* = 1.26 × 10^-2^) (**Figure 4B, Table S12)**.

**Figure 4.**
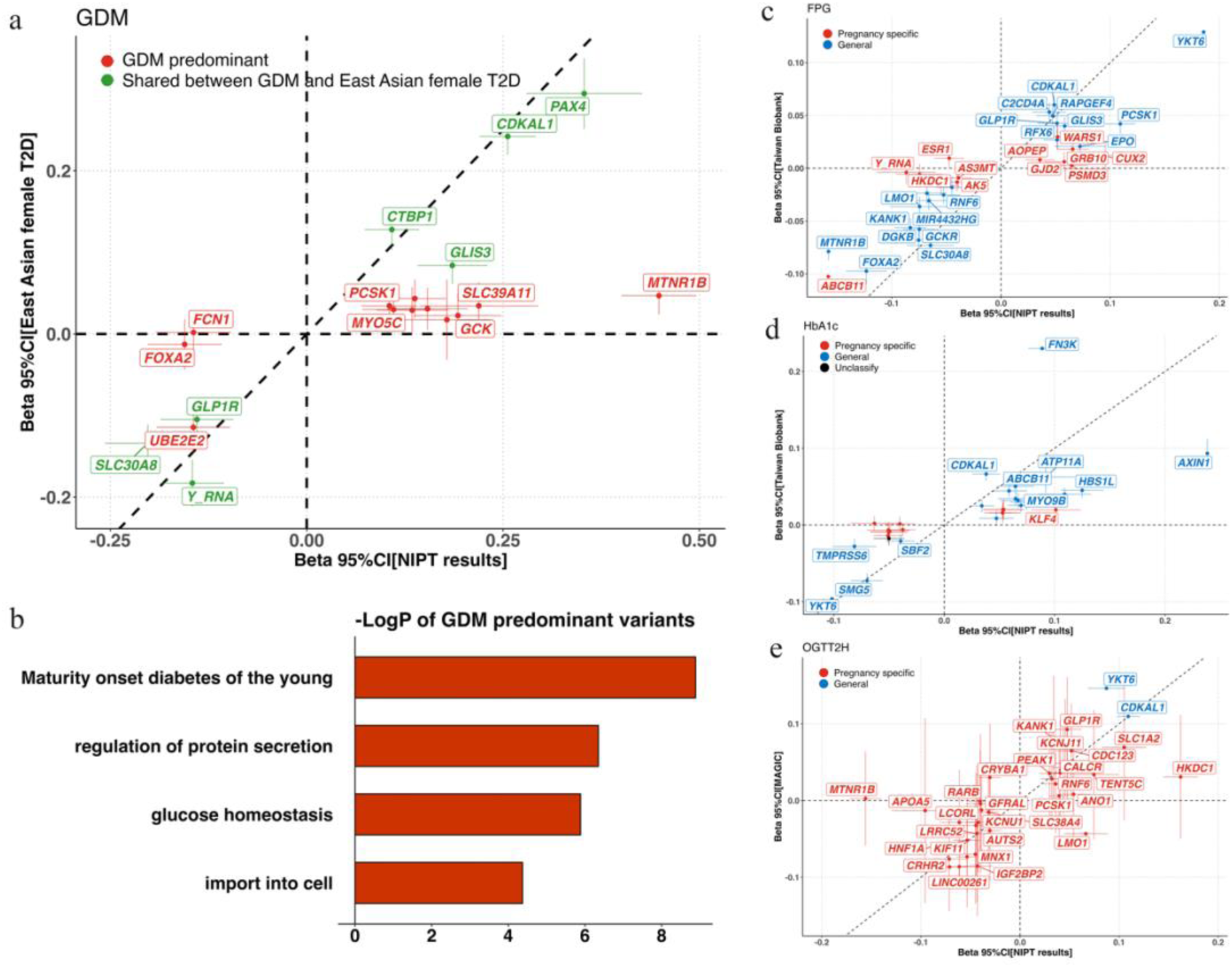
Classification of gestation-specific genetic effects for GDM and three glycemic phenotypes. Panel A: Comparison of the effects of East Asian T2D and clustering results for GDM lead SNPs. Panel B: Enrichment analysis of the two cluster using Metascape 3.5. Only pathways with *P* < 0.05 for the gestation-specific loci are presented in this Figure. Panel C and D: Classification results from the combined ‘linemodels’ and colocalization analyses compared with GWAS data from Taiwan Biobanks for FPG and HbA1c, respectively. Panel E: Results of the ‘linemodels’ analysis compared with GWAS data from the MAGIC consortium for OGTT2H. East Asian T2D data were derived from the East Asian meta-analyses of 23 T2D GWAS, including 27,370 female T2D cases and 135,055 female controls from China, Japan, Korea, Philippines and Malaysia, as part of the Asian Genetic Epidemiology Network (AGEN) and the Diabetes Meta-analysis of Trans-ethnic Association Studies (DIAMANTE) (PMID: 32499647).

### Gestation-specific genetic effects on glycemic traits

To investigate whether genetic effects on glycemic traits differ between pregnant and non-pregnant populations, we applied the same Bayesian classification and colocalization methods (**Figure S13**). Genetic effects on FPG, HbA1c, as estimated in this study, were compared with results from the Taiwan Biobank, while genetic effects on OGTT2H were compared with findings from the MAGIC East Asian population. Genetic loci associated with these glycemic traits were classified into three categories: general, gestation-specific, and unclassified (**Figure 4C-E, Table S5).** For FPG, gestation-specific loci included *AK5, ABCB11, NSD1, Y_RNA, ESR1, GRB10, AOPEP, HKDC1, AS3MT, CUX2, WARS1, GJD2* and *PSMD3*. Gestation-specific loci for HbA1b included *GCKR, PCKS1, LINC00240, WASL, KLF4, CUX2* and *MTCP1*. For OGTT2H, with the exception of *CDKAL1* and *YKT6*, all loci were found to be gestation-specific (**Table S5**). The disproportionately high number of gestation-specific loci for OGTT2H may be attributed to the limited statistical power of OGTT2H GWAS in East Asian populations, as reported by the MAGIC study (**Figure S10**).

### Prediction of GDM in early pregnancy using genetic data

This GWAS elucidates the unique genetic architecture of GDM and glycemic traits in an East Asian population. To explore the predictive potential of integrating genetic information into GDM risk models, we constructed polygenic risk scores (PRS) for GDM, five glycemic traits, and 43 biomarkers (**Table S13**). A tree-boosting machine learning model^23^ incorporating the PRS to the phenotypic data was developed, and Shaley values^24^ was leveraged for interpretability of GDM risk before the 20^th^ gestational week (**Figure 2**).

We used the PRSice-2 algorithm^25^ and PLINK (version 2.0) to construct the PRS models, with details of *P*-value thresholds, SNP counts, and *R^2^* values provided in **Table S14**. Subsequently, we created three decision tree models: (1) a non-genetic model using 46 clinical features before 20 weeks of gestation, (2) a genetic model incorporating 49 PRS values, and (3) a combined model integrating PRS with clinical features. Each of these tree models was trained in 60% of the data from the Baoan 20K cohort, with testing and validation conducted on 20% and the remaining 20% of the cohort, respectively, along with an external validation cohort (NIPT PLUS) (**Figure 2**).

The non-genetic model, based on 46 clinical features, achieved an AUC of 0.712, and an accuracy of 0.835 in the test dataset (**Figure 5A, Table S15**). Key contributors were FPG (measured at ∼16 gestational weeks, 38.12%), maternal age (14.03%), and BMI (measured at ∼16 gestational weeks, 11.39%) (**Figure S14, Table S16**). The genetic PRS model yielded an AUC of 0.587 and an accuracy of 0.805, with PRS for OGTT2H, OGTT1H and GDM being the top three contributors, collectively accounting for more than 80% of GDM liability at 24-28 gestational weeks (**Figure 5A, Figure S14 and Table S17**). The combined model demonstrated the highest predictive performance, with an AUC of 0.729 and an accuracy of 0.835. Key contributors to this model were FPG, maternal age and BMI, followed by the PRS for OGTT2H **(Figure 5A-B, Figure S14 and Table S18)**. In validation cohorts, the combined model achieved AUCs of 0.729 and 0.710 in the Baoan 20K and NIPT PLUS cohorts, respectively, with accuracy rates of 0.841 and 0.806. Compared to the phenotypic model, the combined model showed significance improvement in predictive performance, as demonstrated by DeLong’s test (Baoan 20K validation: *P* = 1.68 × 10⁻²; NIPT PLUS validation: *P* = 4.54 × 10⁻⁵; **Table S19)**.

**Figure 5.**
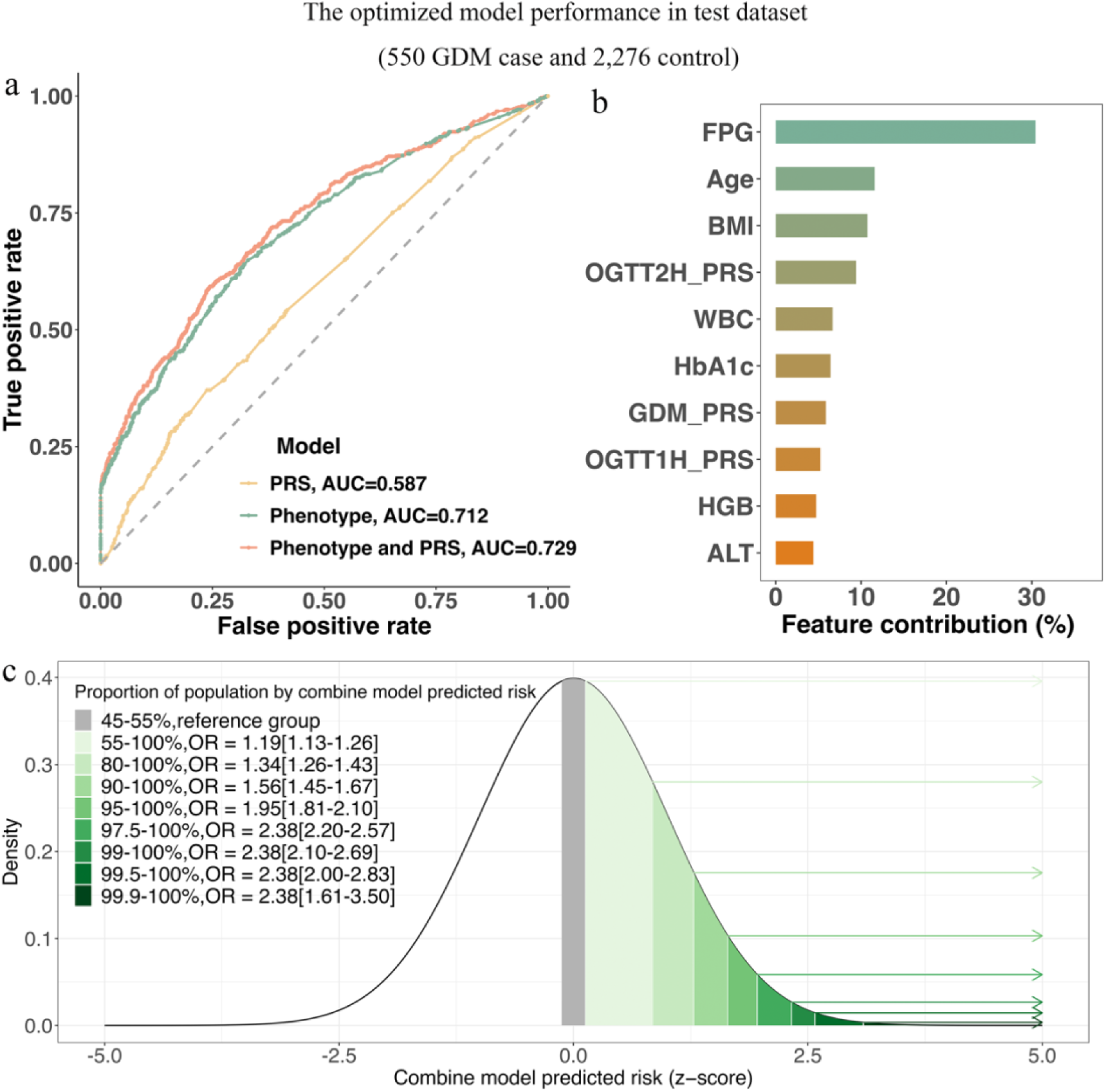
Performance of the GDM predictive model. Panel A: auROC curve for three models. Panel B: Top 15 features of the integrated model, with the x-axis presenting feature contributions based on Shapley values. Panel C: GDM risk at specific percentages of the prediction model.

Shapley values revealed that the majority of features positively correlated with GDM risk **(Figure S14)**. The combine model demonstrated strong predictive power, with individuals in the top 5% of risk scores having a 1.95-fold higher odds of developing GDM (OR = 1.95, 95% CI: 1.81–2.10, *P* < 2.00 × 10^-16^). Those in the top 2.5% had a 2.38-fold higher odds (OR = 2.38, 95% CI: 2.20–2.57, *P* < 2.00 × 10^-16^) **(Figure 5C, Table S20)**. Consistent results were observed in the validation cohorts, with top 2.5% risk scores yielding odds ratio of a 2.31 (95% CI: 2.12–2.52, *P* < 2.00 × 10^-16^) in Baoan 20K and 2.32 (95% CI = 2.15–2.50, *P* < 2.00 × 10^-16^) in NIPT PLUS **(Figure S16, Table S20)**.

All three models demonstrated stable performance, with consistent accuracy rates above 0.8 and AUC fluctuations of around 0.1 across corss-validation splits **(Table S15)**. Calibration plots confirmed the superior alignment between predicted and observed risk in the combined model (**Figure S17**). These findings underscore the values of incorporating genetic data into early GDM prediction models. Notably, the combined model, which relies on clinically accessible NIPT data, offers a cost-effective and practical solution. Moreover, the same training framework can be extended to other non-NIPT datasets using the GWAS summary statistics provided in this study.

## Discussion

This study provides the first comprehensive insights into the genetic architecture of GDM in East Asians and introduces a model that integrates PRS with early electronic health records to enhance the prediction and classification of GDM risk. We conducted the largest GWAS to date on GDM and five glycemic traits in an East Asian cohort, revealing a refined genetic architecture of GDM. Our findings led to the identification of 13 novel loci for GDM and 111 loci for glycemic straits. Notable novel loci for GDM include *PAX4*, a transcription factor crucial for pancreatic islet embryonic development^26^ (lead SNP rs61160304-T, OR[95%CI] = 1.42[1.32-1.53], *P* =3.06⊆10^-21^), and *GLIS3*, which plays a key role in thyroid hormone biosynthesis and pancreatic beta cell function^27^ (lead SNP rs10758593-A, OR[95%CI] = 1.20[1.15-1.26], *P* =9.57⊆10^-17^). Additionally, we identified 12 loci, including eight novel discoveries, that are specific to GDM, revealing distinct genetic mechanisms for GDM compared to T2D in East Asian populations. Our study further highlight 63 potential gestation-specific genetic effect associated with FPG, OGTT2H and HbA1c levels. These findings provide a robust genetic foundation for the development of our GDM prediction model.

Despite the high genetic heritability and polygenic nature of GDM, previous predictive models have generally not incorporated genome-wide genetic data. This gap is likely due to the lack of ancestry-specific GWAS information and the perceived additional cost of genetic testing (**Table S21**). Our model, however, leverages data from widely adopted NIPT data, thus eliminating the need for extra costs. In recent years, NIPT sequencing has revolutionized pregnancy screening worldwide^28^, accumulating a vast amount of genetic data that serves as a valuable resource^12^. In this study, the combined model achieved an AUC of 0.710 and 0.729, and an accuracy of 0.841 and 0.807 for predicting GDM before 20 weeks of gestation in the two validation datasets, significantly outperforming models relying solely on phenotypic data (DeLong’s test *P* < 0.05), which achieved an AUC of 0.710 and 0.681 and an accuracy of 0.815 and 0.777 in the first and second validation datasets, respectively. Our explainable tree-based models identified key predictive factors for GDM, such as gestational age, BMI and FPG levels in early pregnancy. These findings align with previous predictive model based on clinical features^10^, reinforcing the importance of early glucose levels in GDM prediction. Notably, PRS for GDM, OGTT2H and OGTT1H emerged as crucial predictors, highlighting the value of integrating genetic information into GDM prediction models. Compared to earlier studies on early GDM prediction that incorporated genetic data, one study involving 962 participants in China reported an AUC of 0.620 using clinical records and 4 SNPs associated with T2D^11^. Another study, involving 5,085 Chinese participants, achieved an AUC of 0.960 using copy number variants but it lacked external validation^29^**(Table S21)**. Our study demonstrates superior performance, reliability, and robustness in predicting GDM risk.

These findings provide scientific evidence supporting the incorporation of genetic data in GDM prevention and treatment strategies. Moreover, our predictive model could inform a more selective screening process for GDM diagnosis and support early interventions to prevent or mitigate GDM and its associated adverse health outcomes. For individuals identified as at high risk of developing GDM, it is recommended that monitoring be increased and intervention strategies, informed by clinical experience, be implemented as early as medically permissible.

### Limitations of the study

First, the relatively small sample size of the replication cohorts (Baoan 20K and NIPT PLUS cohorts) may lead to an underestimation of the replication rate in our study. To address this limitation, we have made the full GWAS summary statistics publicly available, which will facilitate more accurate estimates of replication rate in future studies. Second, our study highlights the need for further mechanistic investigations into the gestation-specific and shared genetic effects for GDM compared to T2D. Understanding how these genes interact with gestational status and contribute to GDM on the epigenomic, transcriptomic, metabolomic and proteomic levels will provide a more in-depth understanding of GDM and may lead to the development of new therapeutic strategies. Third, the prediction model was evaluated exclusively in an East Asian population, and its applicability to other populations requires further validation. However, we observed a significant genetic correlation for GDM between our study participants and the European population (rg = 0.439 [0.096]), suggesting the potential for generalizability of the model to other populations. With the rapid global accumulation of NIPT sequencing data, future efforts to develop and evaluate prediction models in European populations are warranted. Fourth, previous studies have indicated that copy number variations (CNVs) can serve as predictors of GDM in early gestation^29^. Although CNV analysis was not included in our study, integrating CNVs with PRS in future research may further enhance the model’s predictive performance for GDM. Additionally, we lacked accurate data on behavioral factors, such as personal eating habits, which are associated with the onset of GDM. Incorporating such epidemiological information into the current model may further improve its predictive accuracy. Finally, our method may have limited applicability in hospital settings where NIPT is not routinely offered as a standard screening tool for fetal trisomies. Nevertheless, since the publication of the NEXT RCT study in 2015^30^, NIPT has increasingly been recommended as a first-tier screening tool for trisomy prevention in many countries and is now widely adopted in clinical practice worldwide. The incorporation of genetic data into GDM prevention and treatment strategies could be particularly valuable in these settings.

## Methods

### Participant recruitment

We recruited 121,556 pregnancy participants from Shenzhen Baoan Women’s and Children’s Hospital (referred to as Baoan) and Longgang District Maternity and Child Healthcare Hospital of Shenzhen City (referred to as Longgang). These participants underwent NIPT with an average of 0.17x sequencing depth during the first or second trimester (mean gestational age: 16 weeks) between 2017 and 2021. After excluding 5,412 participants with multiple NIPT sequencing records, 116,144 women remained in the study **(Figure 2)**. The Longgang cohort included 7,438 GDM cases (15.5%) and 40,446 controls, while the Baoan cohort included 4,586 GDM cases (14.4%) and 27,399 controls. Demographic data and measurements for 43 biomarkers collected before 20 weeks of gestation were also obtained (**Table S1**). Furthermore, we collected data from two independent cohort. The first cohort, referred to as the Baoan 20K cohort, included 20,439 participants recruited after 2021 from Shenzhen Baoan Women’s and Children’s Hospital and was not included in the Baoan cohort. After excluding participants without a clear GDM diagnosis or those who underwent NIPT after 20 weeks of gestation, 14,129 participants remained for training and evaluating the prediction model. The second cohort, referred to as the NIPT PLUS cohort, served as an external validation cohort. This cohort included 5,897 individuals who underwent deeper sequencing (0.3x), comprising 795 GDM cases (22.3%) and 2,770 controls with clear GDM diagnosis records before 20 weeks of gestation (detailed information is provided in the Supplemental Notes).

Written informed consent was obtained from all participants. This study was approved by the Ethics Committee of the School of Public Health (Shenzhen), Sun Yat-Sen University (approval number: 2021. No.8), as well as the Institutional Review Boards of Shenzhen Baoan Women’s and Children’s Hospital (approval number: LLSC2021-04-01-10-KS) and Longgang District Maternity and Child Healthcare Hospital (approval number: LGFYYXLLL-2022-024). Data collection was authorized by the Human Genetic Resources Administration of China (HGRAC).

### Phenotype definition

After excluding patients with pre-gestational diabetes mellitus (PGDM), GDM was diagnosed by a 75g oral glucose tolerance test (OGTT) administered between the 24^th^ to 28^th^ weeks of gestation. GDM cases in this study were identified by a clinician based on any of the following criteria: 1) OGTT0H ≥ 5.1 mmol/l (92 mg/dl); 2) OGTT1H ≥ 10.0 mmol/l (180 mg/dl); 3) OGTT2H ≥ 8.5 mmol/l (153 mg/dl) ^31^.

### GWAS with NIPT data

High-quality genotype imputation and accurate genetic effect estimates for GWAS can be achieved using low-depth whole-genome sequencing data generated from NIPT, as demonstrated in our previous and recent studies^12, 13^. The analytical pipeline is available at https://github.com/liusylab/NIPT-human-genetics. Following this protocol, we performed genotype imputation using the Genotype Likelihoods Imputation and Phasing Method (GLIMPSE) software (version 1.1)^32^ utilizing a reference panel of 10,000 Chinese with high-depth sequencing data.

Subsequently, we conducted GWAS using linear or logistic regression models implemented in PLINK (version 2.0)^33^, adjusting for maternal age, BMI, gestational week, and the top 10 principal components as covariates (detailed information is provided in the **Supplemental Notes**). As a secondary analysis, we also performed GWAS excluding BMI as a covariate to assess its impact as a heritable confounder. For meta-analysis, we used fixed-effects models with inverse variance weighting to pool dataset-specific variant effect estimates and their standard errors using METAL (version 2011-03-25)^34^. To identify independent genome-wide significant signals, we employed the genome-wide complex trait analysis-conditional and joint association analysis (GCTA COJO)^35^ with stepwise model selection (–cojo-slct) and a collinearity threshold of 0.2, filtering out variants with a minor allele frequency (MAF) < 0.01. The lead SNP was defined as the variant with the smallest p-value within a 500kb region upstream and downstream, and the locus was defined as the 1Mbp region centered on the lead SNP.

We also compared the GWAS results obtained using PLINK2 with those from REGENIE (v4.1) ^36^and BOLT-LMM (v2.4.1)^37^, which demonstrated high consistency across methods. This consistency may be attributed to the absence of cryptic family relatedness in the dataset. Since PLINK2 produced the expected intercepts and ratios for the highest proportions of phenotypes, we selected it as the primary method for reporting results.

### Definition of novel loci

Independent loci consisting of SNPs located within 500 kb upstream or downstream of a lead SNP previously associated with the same phenotype (i.e., GDM or any of the five glycemic traits) were classified as known loci, while all others were considered novel loci. Previous genetic association findings were determined based on the lastest version of the GWAS Catalog, released on February 18, 2025 (gwas_catalog_v1.0.2-associations_e113_r2025-02-18.tsv).

Specifically, for GDM, novel loci were defined as those that did not contain any SNPs previously reported in genetic association studies within a 1Mbp window centering the lead SNP. We did not consider comparisons with T2D in defining novel loci for GDM.

For glycemic traits, novel loci were defined as those not previously associated with the same glycemic trait, regardless of whether the association was identified in pregnant or non-pregnant individuals. Comparisons with T2D or GDM were not considered in defining novel loci for glycemic traits.

### Replication

To evaluate and replicate the GWAS meta-analysis results for GDM and five glycemic traits, we first compared the genetic effect estimates of the lead SNPs between the separate GWAS conducted at the two hospitals and the meta-analysis results. For external replication, we performed replication analyses in two independent cohorts (Baoan 20K and NIPT PLUS) and compared the meta-analysis results of these cohorts. Additionally, we compared the GWAS meta-analysis results for FPG and HbA1c with summary statistics from the Taiwan Biobank^38^, and for OGTT2H with summary statistics from the MAGIC consortium’s East Asian population^20^. The GDM GWAS findings were further replicated using data from a previous European study^18^. Finally, genetic correlations were estimated using the linkage disequilibrium (LD) score regression, as applied in heritability calculations^9^.

### GDM-specific and shared variants with T2D

We utilized the ‘linemodels’ package to analyze the lead variants identified in the GWAS analyses^22^. The variants were classified into two categories based on their bivariate effect sizes, which were modeled using linear models. Membership probabilities for each variant in the two categories were calculated, assuming equal prior probabilities. Variants were assigned to a specific category if their posterior probability exceeded 0.95. Detailed parameters of the models are provided in the Supplemental Notes. To minimize misclassification due to LD, we further conducted colocalization analysis using the’coloc’ package^39^, to assess whether the genetic loci significantly associated with both phenotypes were the same. Finally, we integrated the results from these two analyses to classify the genetic loci (**Figure S13**).

### Pathway enrichment analysis

Enrichment analysis was performed using the Metascape platform (https://metascape.org/)^40^. This method calculates the hit rate of the genes in our list relative to a background hit rate, with the enrichment factor defined as the ratio of these two rates. The p-value measures the probability of observing multiple pathways, calculated using a cumulative hypergeometric distribution. A more negative p-value indicates a lower likelihood that the observed enrichment is due to chance.

### Development of polygenic risk scores

We employed PRSice-2 (version 2.3.3)^25^ to select SNPs for PRS, utilizing the meta-GWAS analysis results as the base data, which provided the association effect sizes for over 12 million SNPs. The genotype data from the Baoan 20K training dataset (comprising 60% of the Baoan 20K cohort) served as the target data, with phenotypes used to identify the optimal SNPs for the PRS model. During this process, genotype data were pruned using the following parameters: --clump-kb 500, --clump-r2 0.2, ensuring that the LD between SNPs included in the PRS was below 0.2. Finally, we applied PLINK (version 2.0) to calculate the PRS using the selected SNPs (**Figure 2**).

### Development of the GDM early prediction model

We developed three models, each incorporating different features: (1) a non-genetic model based on clinical information collected before 20 weeks of gestation, (2) a genetic model utilizing PRS derived from the NIPT GWAS of GDM, glycemic traits and 43 biomarkers, and (3) a combined model that integrates both PRS and non-genetic factors. In the Baoan 20K cohort, 60% of the participants (1,501 cases and 6,976 controls) were used for training data, with five-fold cross-validation applied.

Randomized splits were performed 10 times for training and testing. GDM recurrence predictions were conducted using XGBoost^41^, with optimal parameters determined through Optuna, a hyperparameter optimization framework^42^. The parameters identified in each round of five-fold cross-validation were then applied to the test set (20% of the Baoan 20K cohort). The model with the best performance in test set was selected as the final model for reporting and validation in the validation set (20% of the Baoan 20K cohort) and external validation cohort (NIPT PLUS). Finally, we employed Shapley Additive Explanation (SHAP) value to interpret the model^43^**(Figure2)**.

## Data availability

The complete GWAS summary statistics for GDM, FPG, OGTT0H, OGTT1H, OGTT2H, and HbA1c have been deposited in the GWAS Catalog database (https://www.ebi.ac.uk/gwas/) and the National Genomics Data Center (NGDC) (https://ngdc.cncb.ac.cn/gvm/), with approval from the China’s National Health Commission (permission number: 2024BAT01079). These data will be publicly available upon publication.

Raw sequencing data have been deposited in the Genome Sequence Archive (GSA) for Humans at the National Genomics Data Center under the BioProject accession number GSA-Human: HRA006833, with approval from China’s National Health Commission (permission number: 2024BAT01079). Access to these data can be obtained through formal applications, following the GSA guidelines (https://ngdc.cncb.ac.cn/gsa-human/document). Data access is restricted to academic research purposes only.

## Code availability

The model depicted in Figure 2 has been submitted to Code Ocean and will be made publicly accessible on GitHub upon publication. The code for performing GWAS on GDM and glycemic traits using non-invasive prenatal test data is available at https://github.com/liusylab/NIPT-human-genetics.

## Data Availability

The complete GWAS summary statistics for GDM, FPG, OGTT0H, OGTT1H, OGTT2H, and HbA1c have been deposited in the GWAS Catalog database (https://www.ebi.ac.uk/gwas/) and the National Genomics Data Center (NGDC) (https://ngdc.cncb.ac.cn/gvm/), with approval from the China National Health Commission (permission number: 2024BAT01079). These data will be publicly available upon publication.
Raw sequencing data have been deposited in the Genome Sequence Archive (GSA) for Humans at the National Genomics Data Center under the BioProject accession number GSA-Human: HRA006833, with approval from China National Health Commission (permission number: 2024BAT01079). Access to these data can be obtained through formal applications, following the GSA guidelines (https://ngdc.cncb.ac.cn/gsa-human/document). Data access is restricted to academic research purposes only.

## Acknowledgments

The study was supported by the National Natural Science Foundation of China (32470642, 82203291 and 31900487), the Shenzhen Science and Technology Program (20220818100717002 and ZDSYS20230626091203007), the Guangdong Basic and Applied Basic Research Foundation (2022B1515120080), and the Shenzhen Health Elite Talent Training Project. The computation was supported by the BrightWing High-performance Computing Platform of School of Public Health (Shenzhen), the High-performance Computing Public Platform (Shenzhen Campus) of SUN YAT-SEN UNIVERSITY), and the National Supercomputing Center in Guangzhou.

## Author Contributors

SL, YG, JZ, and FW conceived and designed the study. YG, HZ and PW formally analysed the data. YG, HZ, PW, XG, YW, ZY and YC performed the visualisation of all results. YG, YL, SC, LH, and XC conducted data pre-processing and the preliminary analyses. JZ, QZ and FW provided the validation data. SL and GC provided professional guidance and interpretation of data. YG and SL drafted the manuscript. All authors acquired and interpreted the data, critically revised the paper, and had final responsibility for the decision to submit for publication.

## Competing interests

The authors declare no competing interests.

## Notes

### Competing Interest Statement

The authors have declared no competing interest.

### Funding Statement

The Guangdong Basic and Applied Basic Research Foundation (2022B1515120080 and 2020A1515110859), the Shenzhen Science and Technology Program (20220818100717002), the Shenzhen Health Elite Talent Training Project, the Economic and Technological Development Special Fundation of Shenzhen Longgang District (LGKCYLWS2022008), the National Natural Science Foundation of China (31900487 and 82203291),National Natural Science Foundation of China (82203291),Shenzhen Health Elite Talent Training Project,the Economic and Technological Development Special Fundation of Shenzhen Longgang District （LGKCYLWS2022008）

### Author Declarations

This study was approved by the Ethics Committee of the School of Public Health (Shenzhen), Sun Yat-Sen University (approval number: 2021. No.8), as well as the Institutional Review Boards of Shenzhen Baoan Women and Children Hospital (approval number: LLSC2021-04-01-10-KS) and Longgang District Maternity and Child Healthcare Hospital (approval number: LGFYYXLLL-2022-024). Data collection was authorized by the Human Genetic Resources Administration of China (HGRAC).

