## Supplemental Figures for "Genetic Architecture and Risk Prediction of Gestational Diabetes Mellitus in over 116,144 Chinese Pregnancies"

*: Those authors contribute equally

#: Correspondence should be addressed to

Siyang Liu or Jianxin Zhen or Fengxiang Wei

### Supplementary Notes

#### Details on the 43 pregnant measurements, GDM and glycemic traits

This study collected 43 pregnant measurements categorized into six biological categories: blood routine (N=24), blood anemia (N=2), kidney function (N=3), liver function (N=8), thyroid function (N=3), and basic information (N=3) **(Table S1)**. For GDM and the five glycemic traits, we conducted the following GWAS: (1) GDM case (N=12,024) versus non-diabetes control (N=67,845); (2) Linear regression on FPG (N=56,912), OGTT0H (N=85,086), OGTT1H (N=85,856) and OGTT2H (N=84,743) and HbA1c (N=69,269).

#### Study and replication population

In addition to the previously mentioned 121,556 pregnancy participants, we collected data from two independent cohort. The first cohort includes 20,439 participants, who sought maternal check-ups at Shenzhen Baoan Women's and Children's Hospital (Shenzhen, China) throughout the entire 40-week gestational period starting from the year of 2022 (referred to as Baoan 20K). After excluding participants without a clear GDM diagnosis or NIPT performed after 20 week of gestation, 14,129 individuals (2,574 cases and 11,555 controls) were included for training and evaluating the prediction model.

The second cohort, used as an external validation cohort, included 5,897 individuals who sought maternity check-ups at Shenzhen Baoan Women’s and Children’s Hospital (Shenzhen, China) throughout their entire gestational period. These individuals underwent NIPT in either the first or second trimester between 2020 and 2021. The samples from this cohort are characterized by a deeper sequencing depth, with an average depth of 0.3x, in comparison to conventional NIPT with an average depth of 0.17x. Ths cohort is referred to as NIPT PLUS cohort.

This study was approved by the Medical Ethics Committee of the School of Public Health (Shenzhen), Sun Yat-sen University, Longgang District Maternity and Child Healthcare Hospital of Shenzhen City, and Shenzhen Baoan Women's and Children's Hospital. Data collection was approved by the Human Genetic Resources Administration of China (HGRAC).

#### GWAS

For the GWAS of GDM and five quantitative glycemic traits GWAS, gestational week, maternal age, BMI and the top ten principal components to account for population stratification were included as covariates. We also conducted a secondary analysis excluding BMI as covariate to account for the heritable confounding. In the GWAS of the 43 pregnancy-related measurements, gestational week was not included as a covariate, whereas BMI, maternal age, and the top ten principal components were retained as covariates.

#### Distinct and shared variants analysis

To address potential power differences between the T2D and GDM GWAS, we employed the ‘linemodels’ and ‘coloc’ approaches. The ‘linemodels’ R package (PMID: 36864614) probabilistically clusters explanatory variables based on effect sizes across multiple outcomes, incorporating scale, slopes, and correlation to account for sample size variability. Scale parameters were optimized using an empirical Bayes method to mitigate the impact of sample size differences on effect size estimates.

In one model, the slope was fixed at 0, while in the other, the model was estimated using an expectation-maximization (EM) algorithm. The scale parameters representing effect size magnitudes were also estimated using the EM algorithm, with correlation parameters set to 0.995 to allow for deviations from the lines. Given that the two GWAS datasets consisted of non-overlapping samples, the correlation between their effect estimators was set to 0.

Furthermore, to assess shared genetic effects across datasets, we conducted a colocalization analysis using the R ‘coloc’ package. This method evaluates the overlap of individual SNPs in high linkage disequilibrium (LD) between datasets using Bayesian testing. Together, these methods minimize the impact of power differences and enhance the robustness of our findings.

### Supplementary Figures

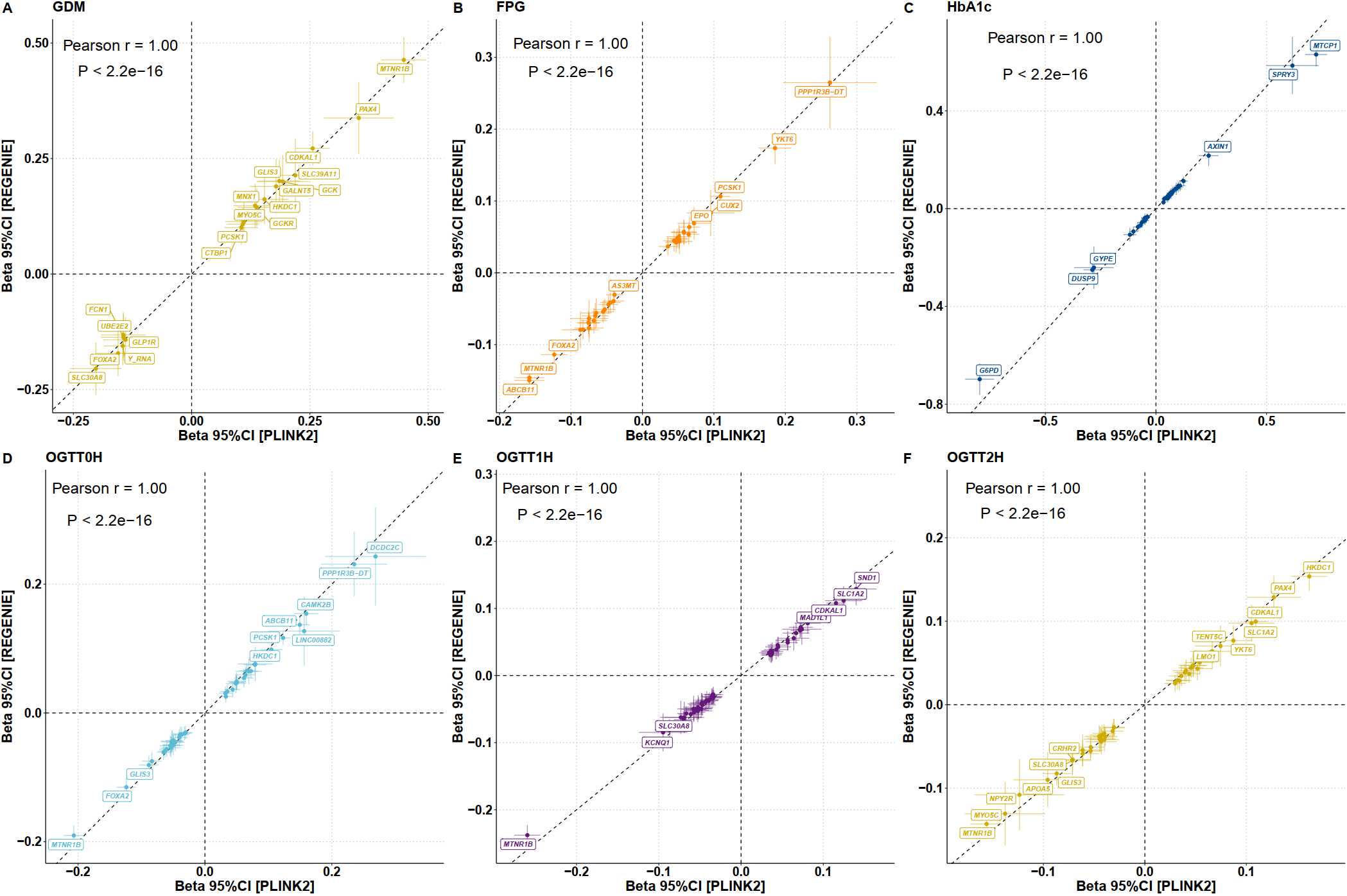

#### Figure S1. Comparison of associations for lead SNPs in GDM and five glycemic traits between PLINK2 and REGENIE.

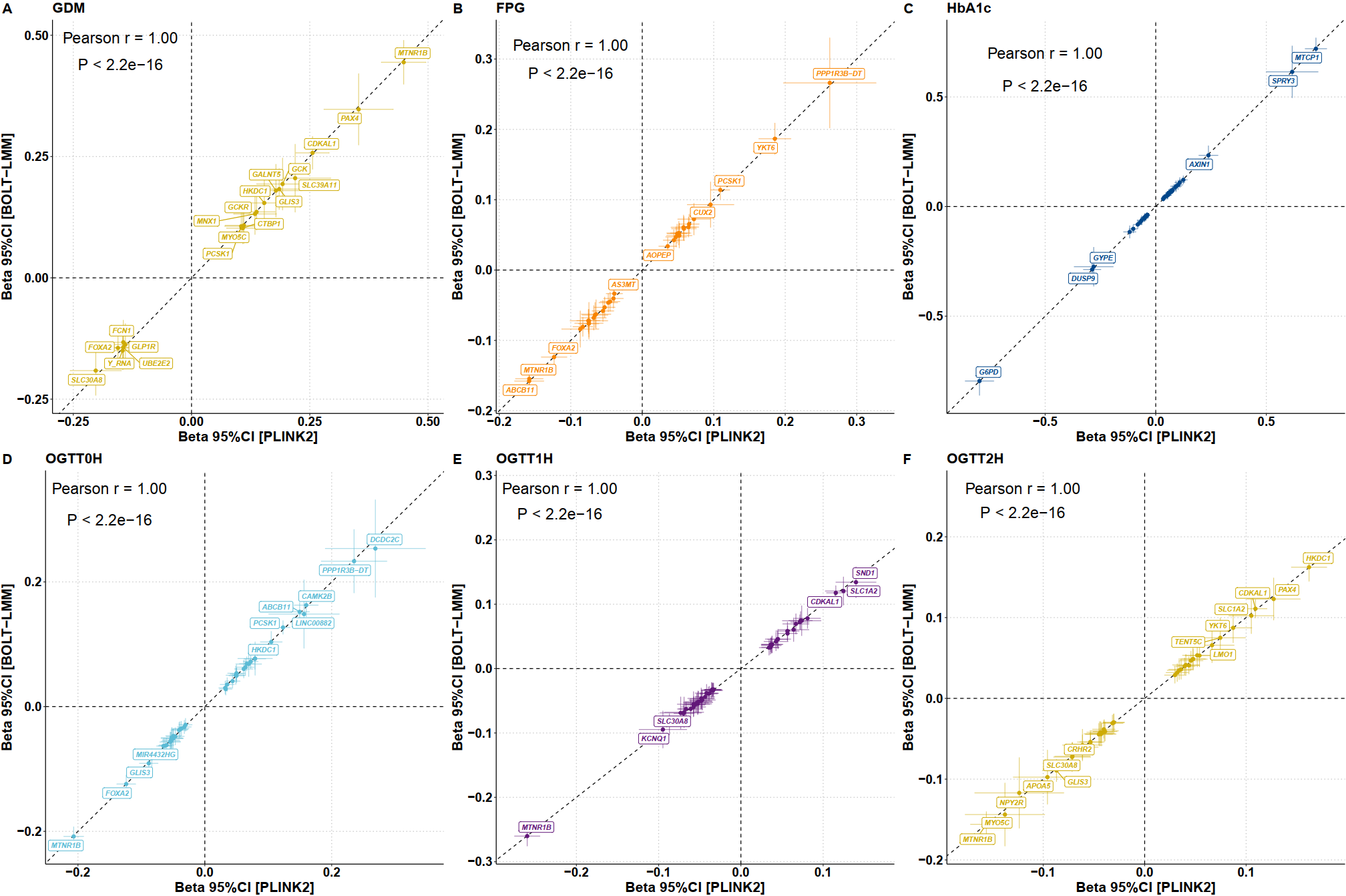

#### Figure S2. Comparison of associations for lead SNPs in GDM and five glycemic traits between PLINK2 and BOLT-LMM.

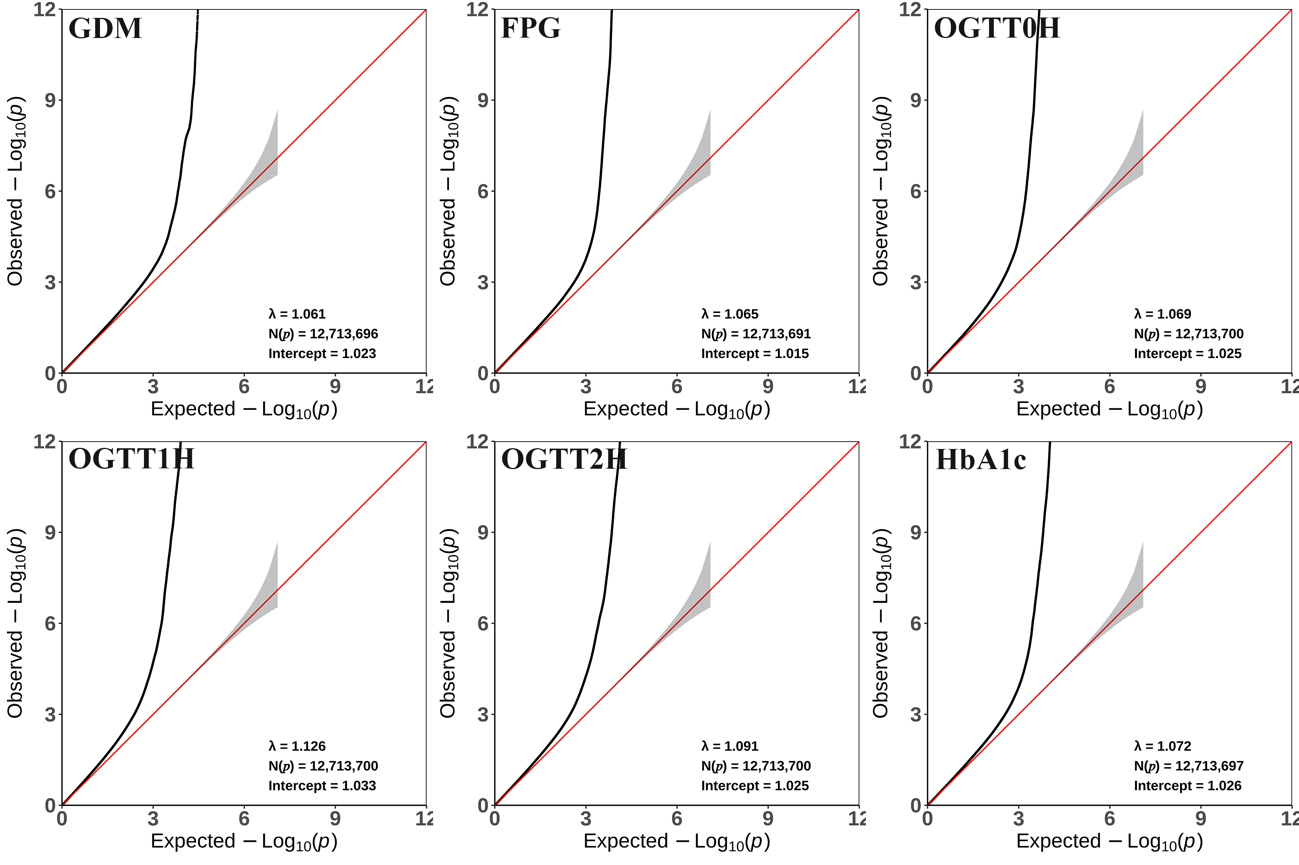

#### Figure S3. Quantile-Quantile (QQ) plot of PLINK2 GWAS for GDM and five glycemic traits.

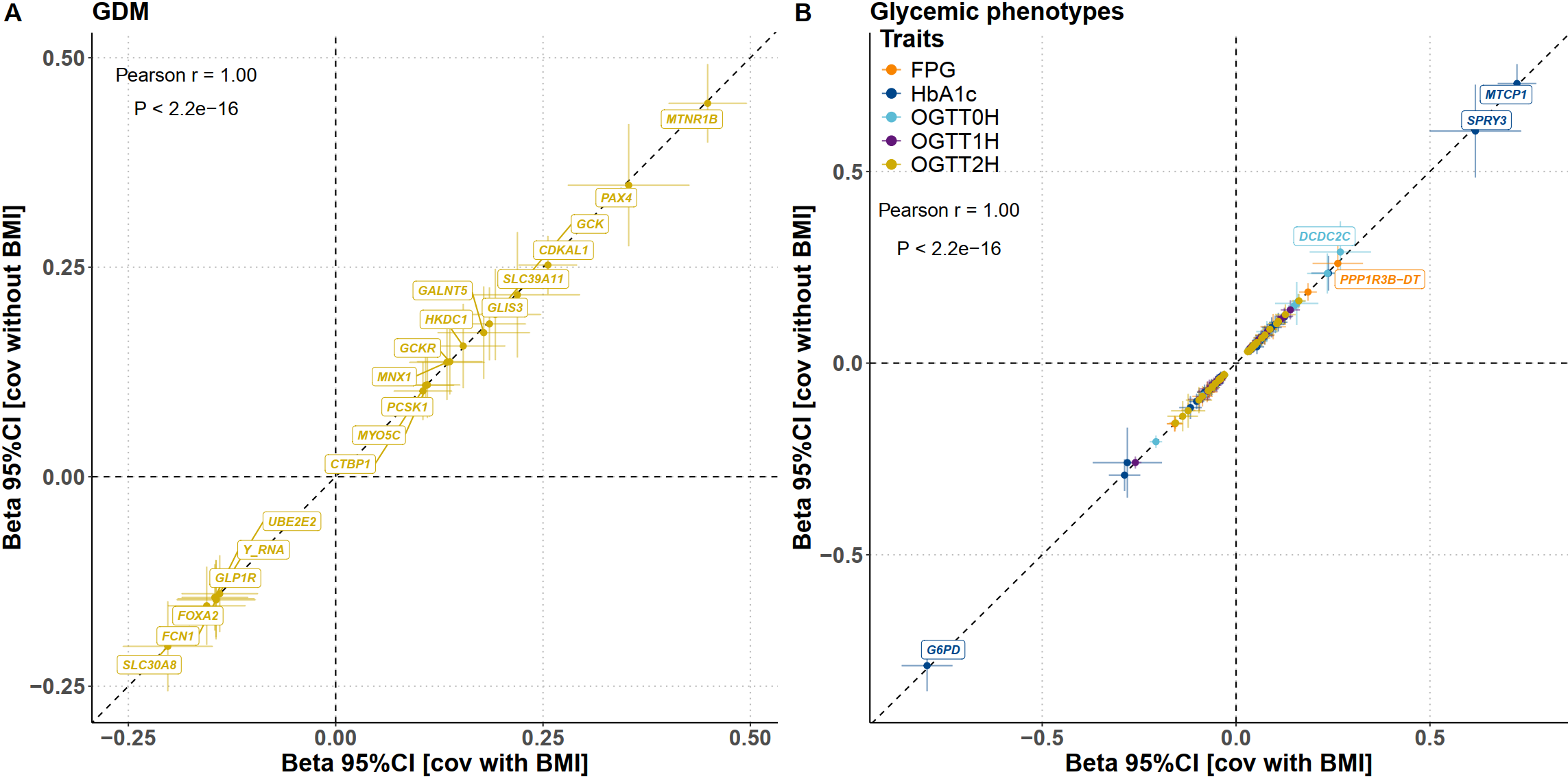

#### Figure S4. Comparison of lead SNP effects between the primary GWAS and secondary analysis excluding BMI as a heritable confounder.

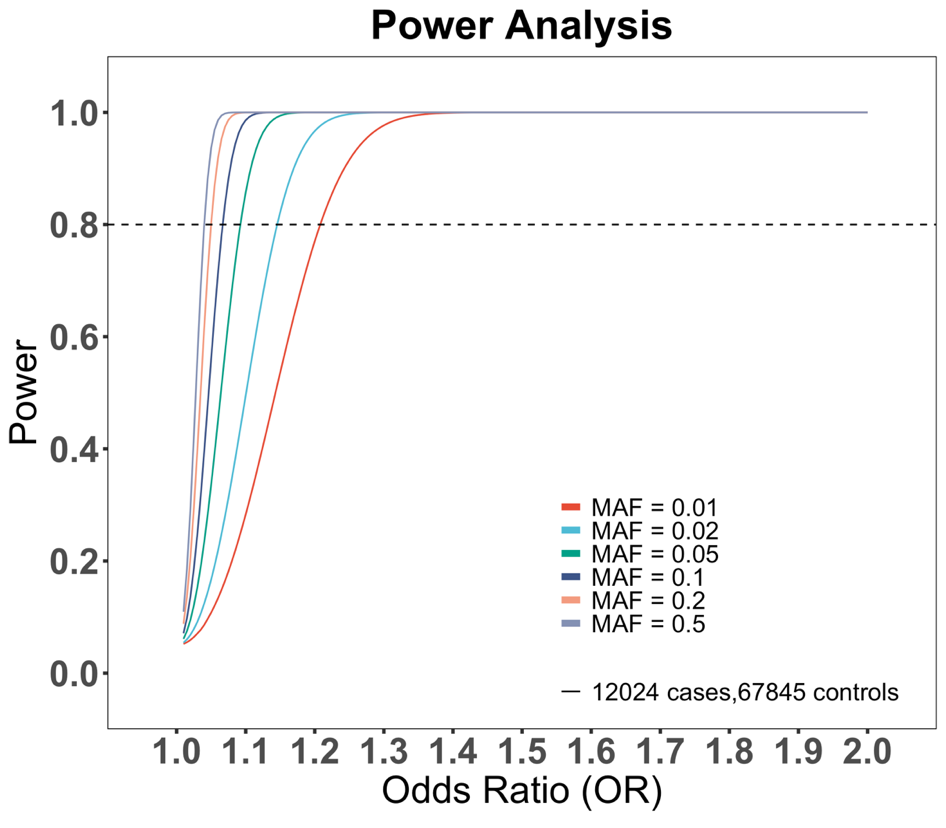

A

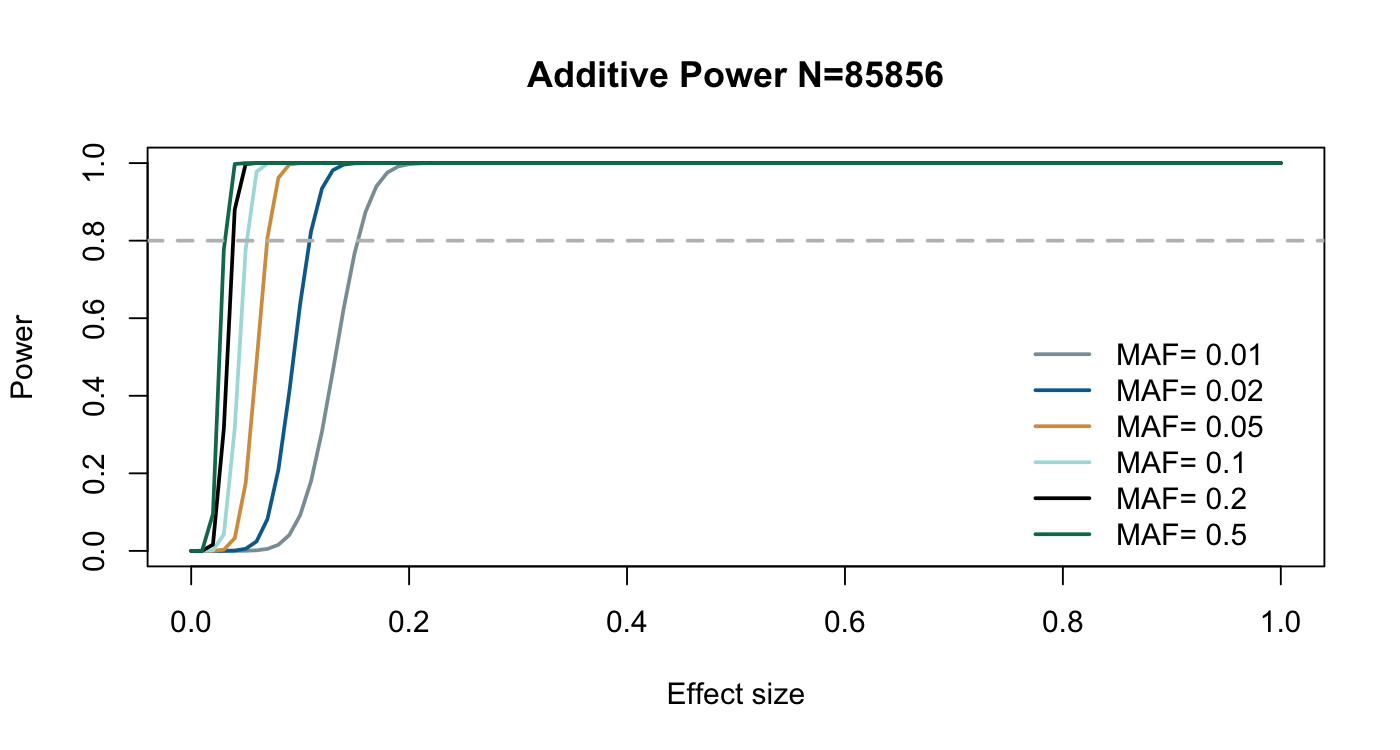

B

#### Figure S5. Power analysis of the genome-wide association study (GWAS).

A: power analysis for GWAS of GDM susceptibility (12,024 cases vs. 67,845 controls) with a significance threshold of *p* < 5🞨10^-8^. B: power analysis for GWAS of quantitative glycemic traits, with a significance threshold of *p* < 5🞨10^-8^. MAF: minor allele frequency. The analysis showed that loci with a minimum MAF of 0.01 and odds ratio (OR) of 1.2, MAF of 0.05 and OR of 1.1, or MAF of 0.2 and OR of 1.01 could be identified for GDM, and a minimum MAF of 0.01 and effect size of 0.15 for five glycemic traits.

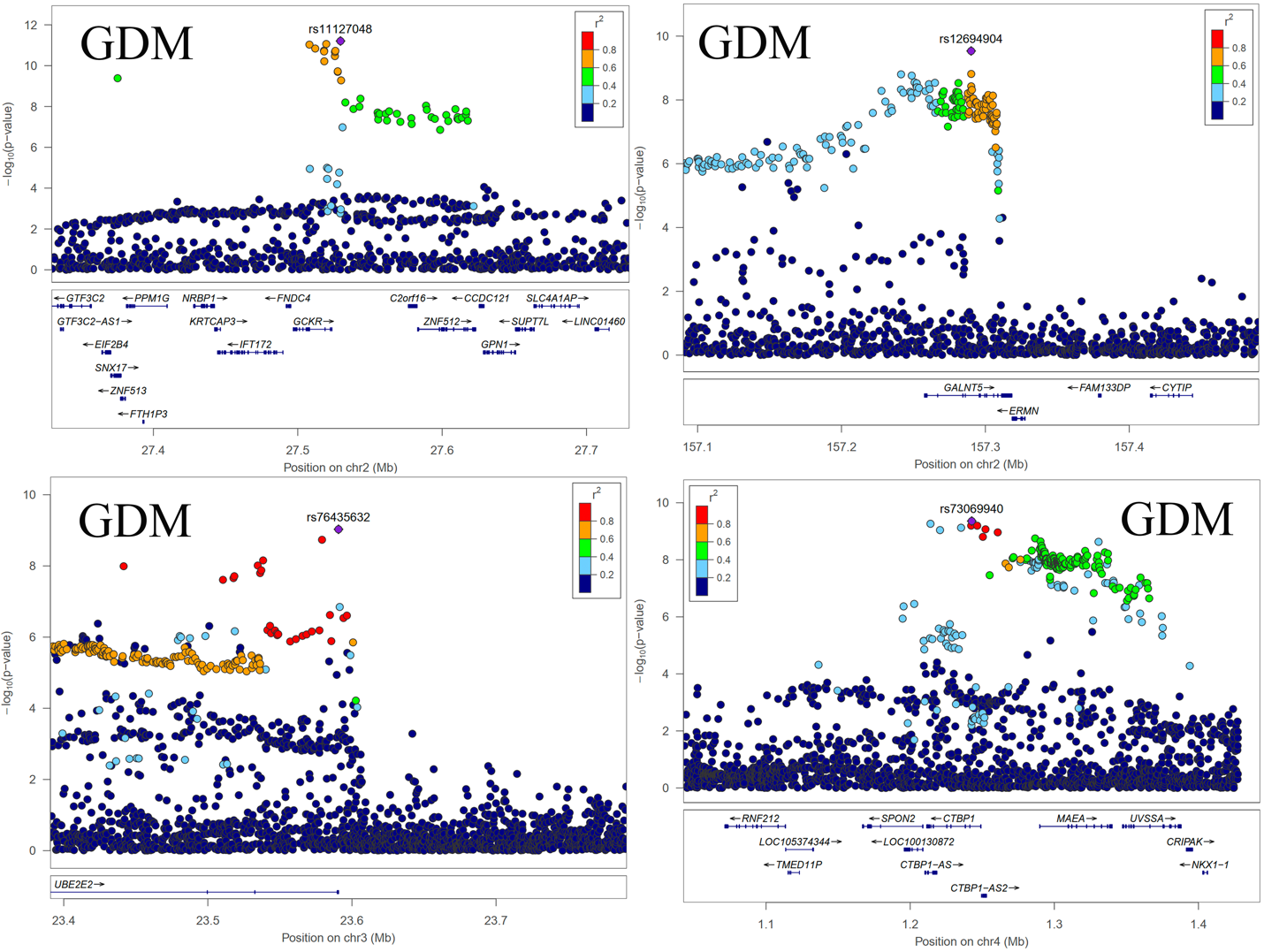

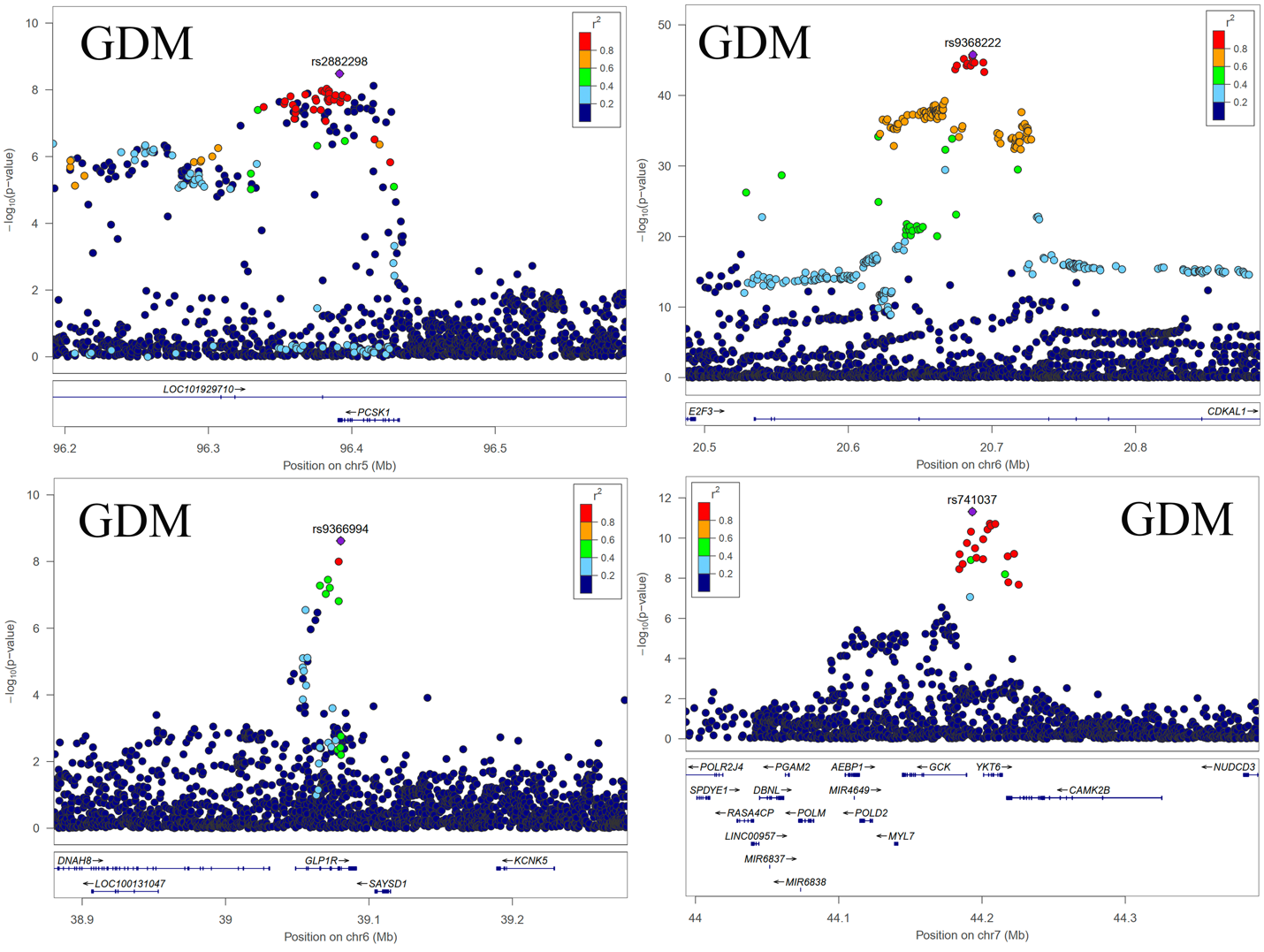

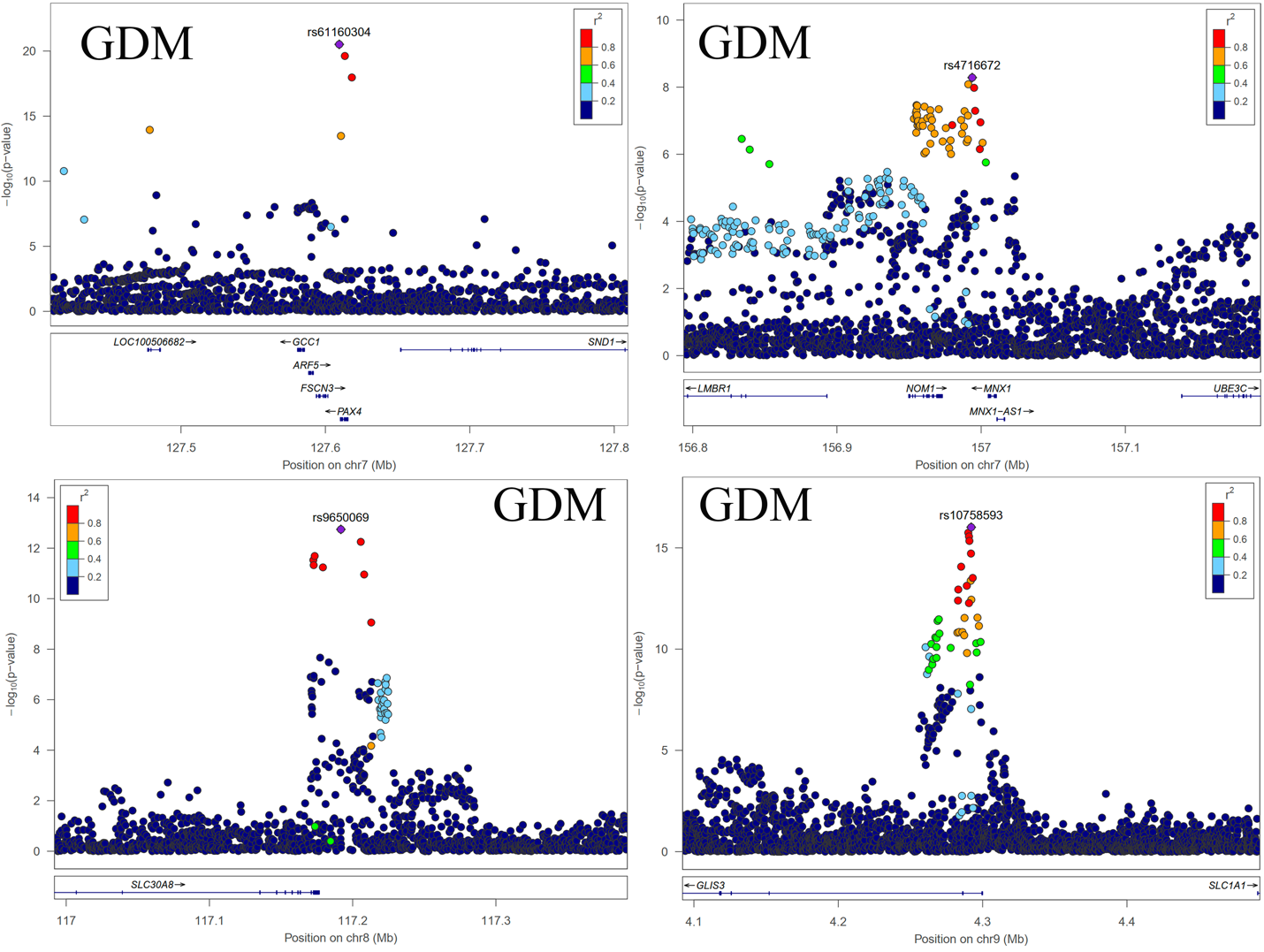

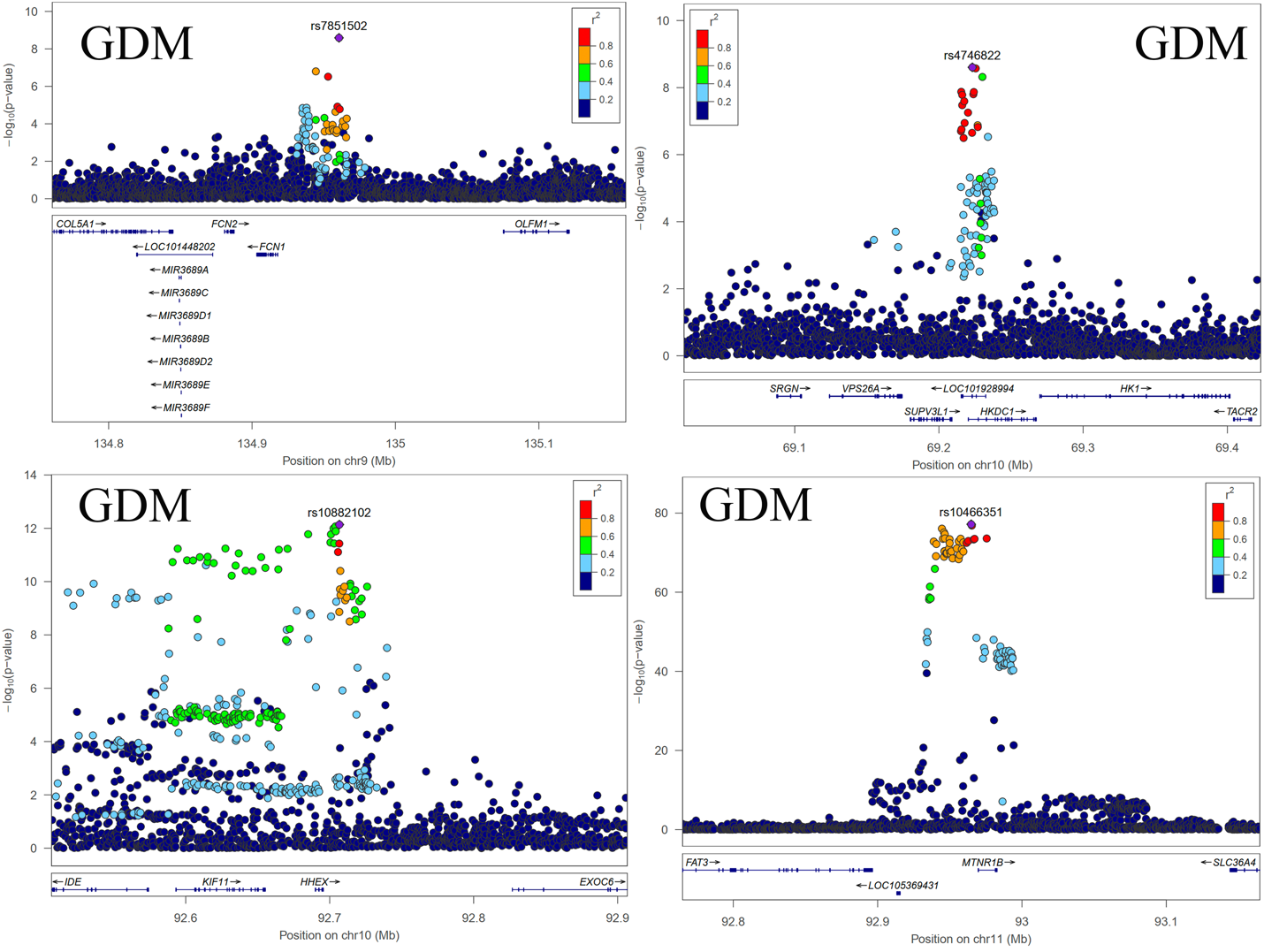

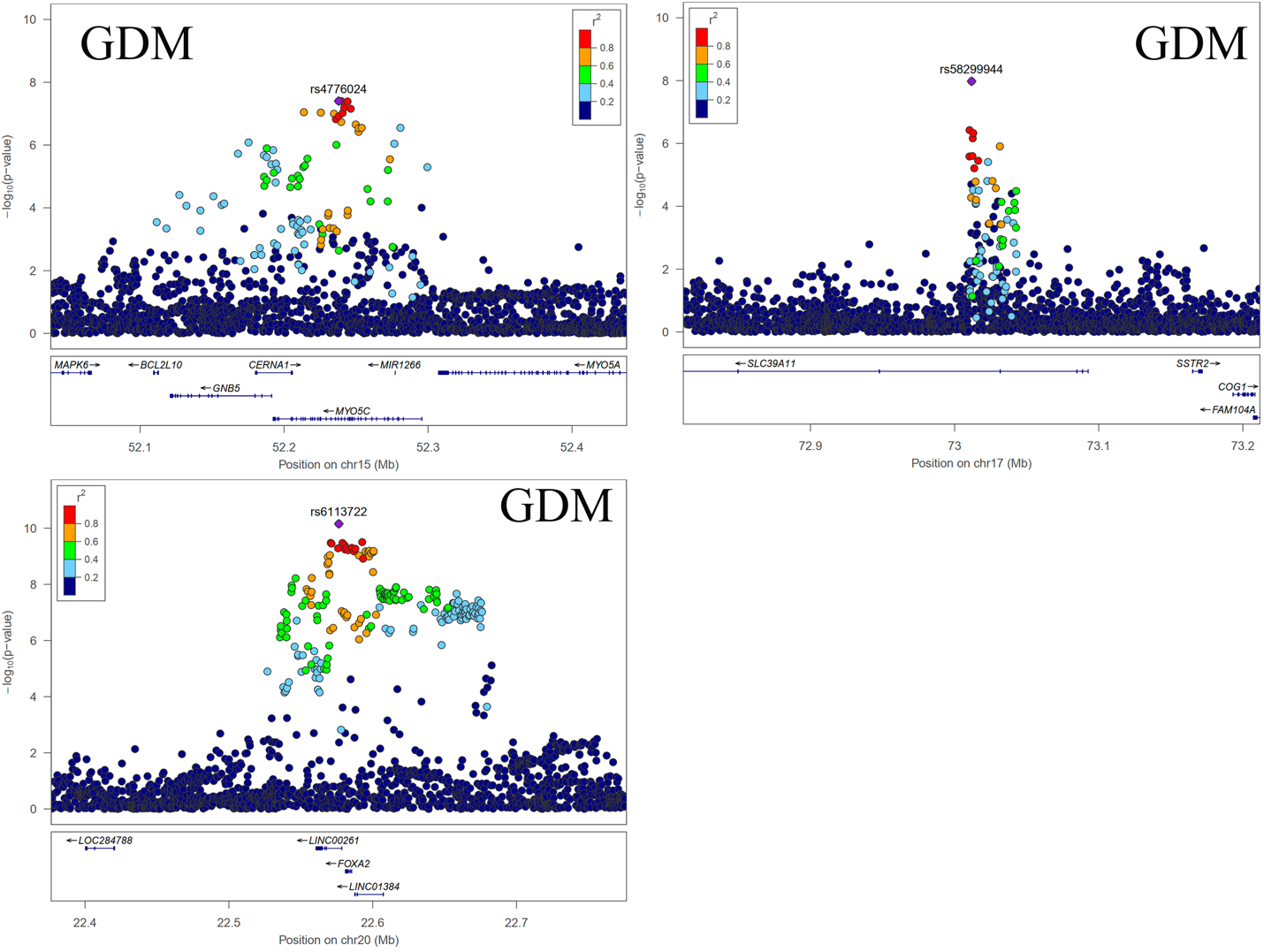

#### Figure S6. LocusZoom plot of genome-wide significant loci associated with GDM in this study.

A regional plot of all 19 lead SNPs for GDM (Table S4) was generated using LocusZoom software, displaying P-values and linkage disequilibrium (LD) (r²) for SNPs within a 500 kb flanking region upstream and downstream.

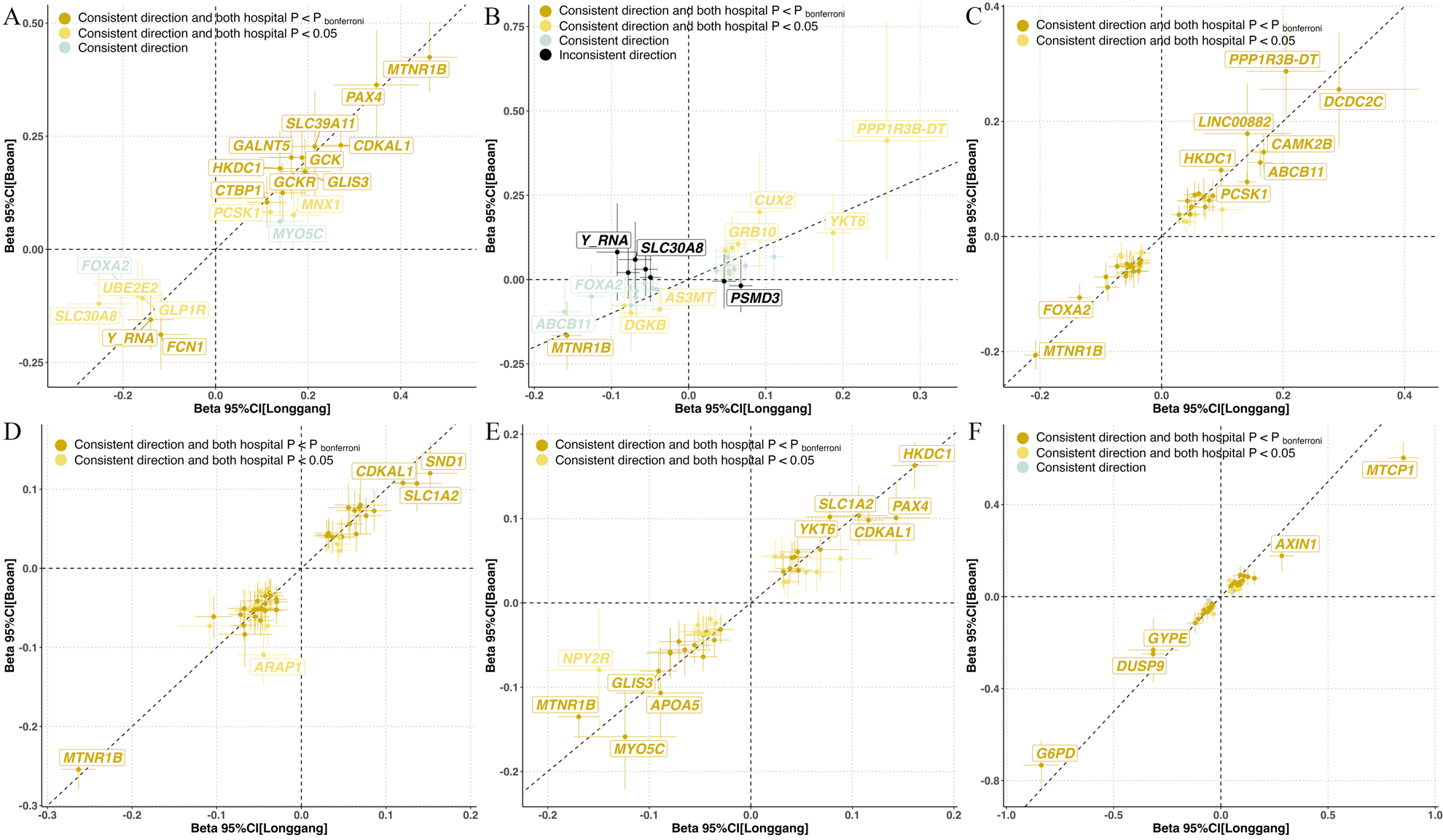

#### Figure S7. Comparison of lead SNPs effects between Baoan and Longgang hospital cohorts.

Panel A-F present the effect comparision of lead SNPs between Baoan and Longgang hospital cohorts for GDM, FPG, OGTT0H, OGTT1H, OGTT2H and HbA1c, respectively.

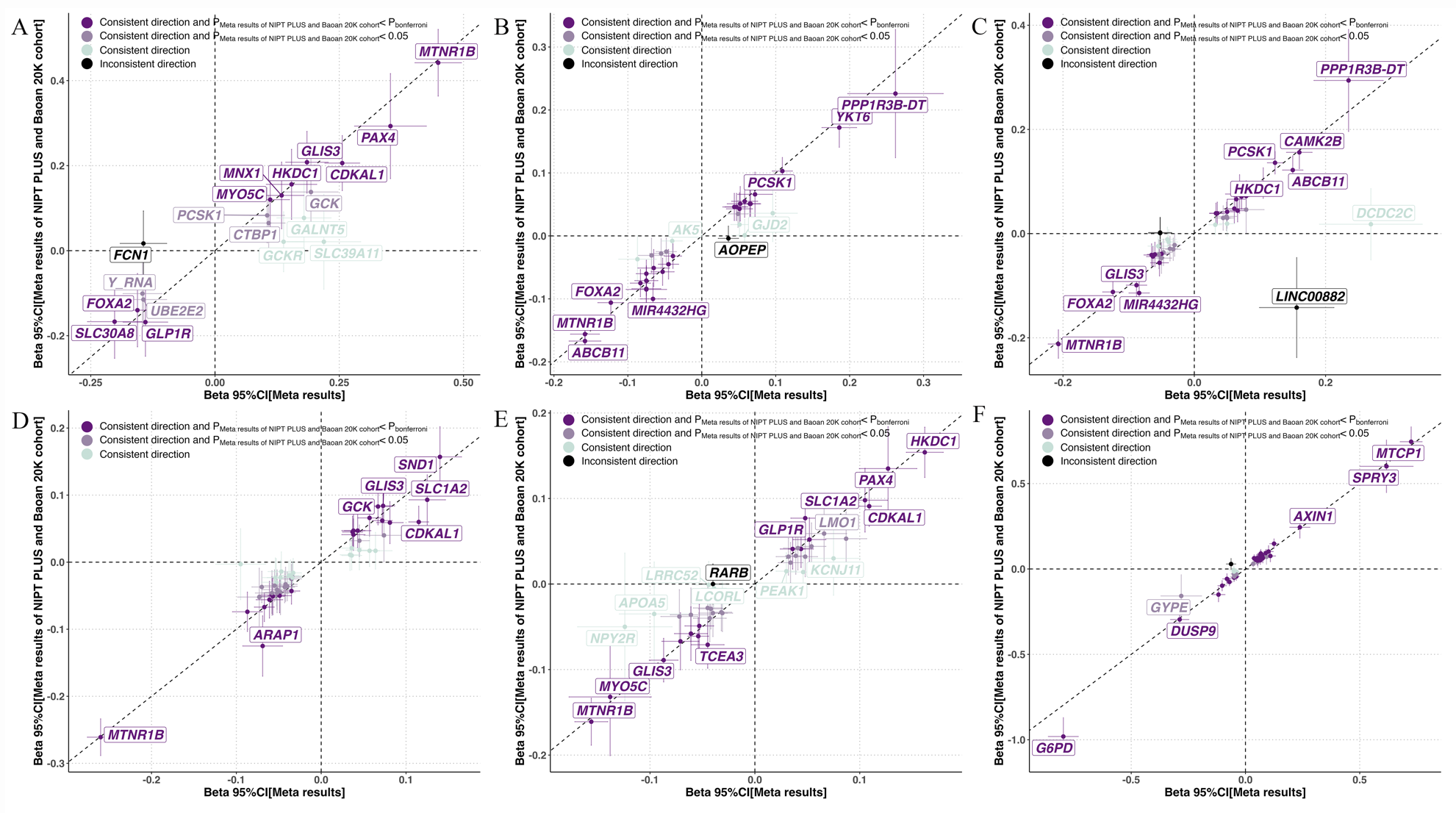

#### Figure S8. Replication of lead SNPs with meta-analysis results from Baoan 20K and NIPT PLUS cohorts.

Panel A-F show the replication of lead SNPs based on meta-analysis results from Baoan 20K and NIPT PLUS cohorts for GDM, FPG, OGTT0H, OGTT1H, OGTT2H and HbA1c, respectively.

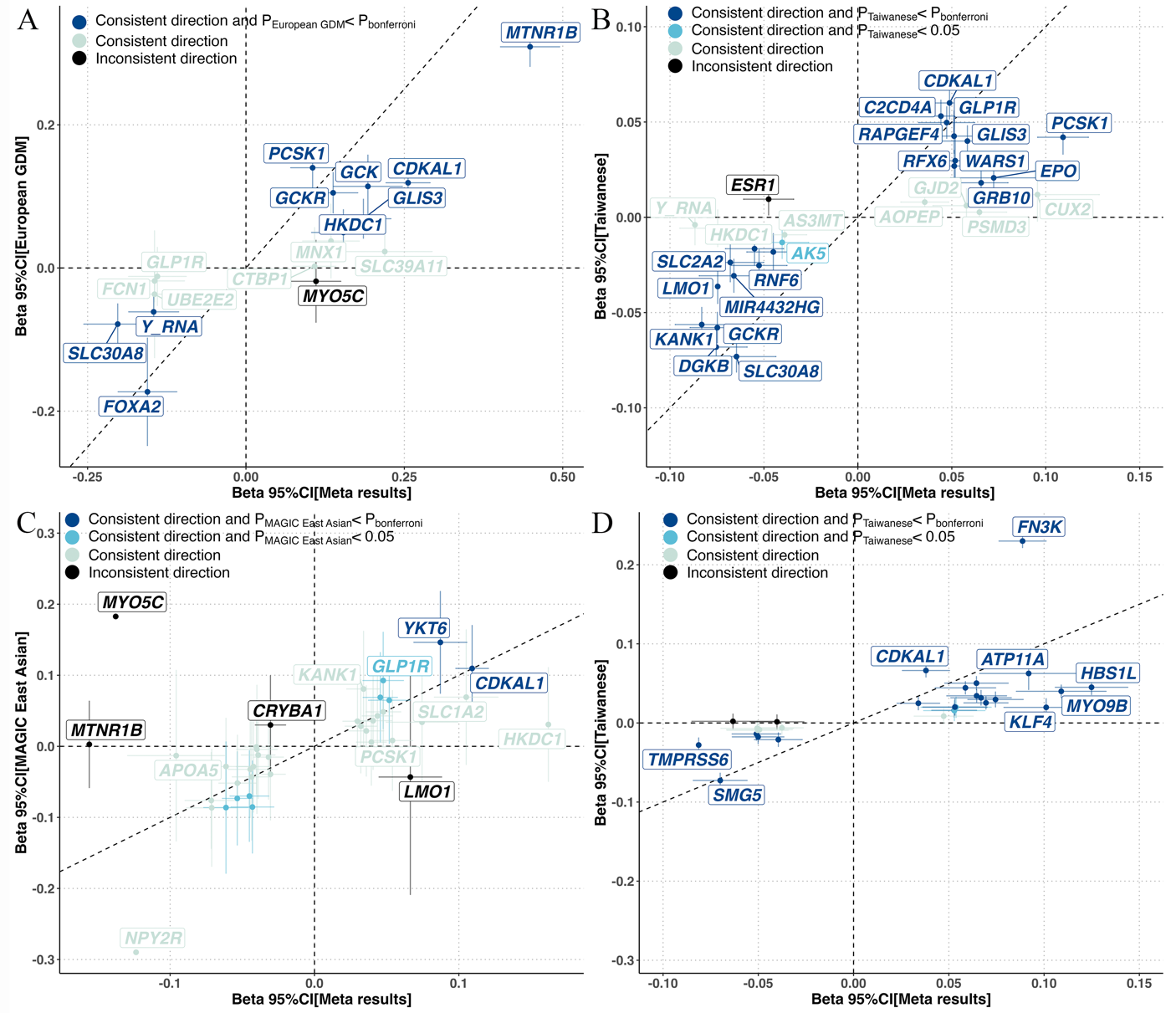

#### Figure S9. Comparison of lead SNP effects with previous studies.

A: Comparison of GDM lead SNPs with data from a European GDM study;

B: Comparison of FPG lead SNPs with data from the Taiwan Biobank study;

C: Comparison of OGTT2H lead SNPs with data from the MAGIC East Asian study;

D: Comparison of HbA1c lead SNPs with data from the Taiwan Biobank study.

Summary statistics for the European GDM study were obtained from the FinnGen study (GCST90296696). The OGTT2H GWAS summary statistics for East Asian populations were downloaded from the MAGIC consortium (GCST90002226). Summary statistics for FPG (GCST90278628) and HbA1c (GCST90278632) were obtained from the Taiwan Biobank.

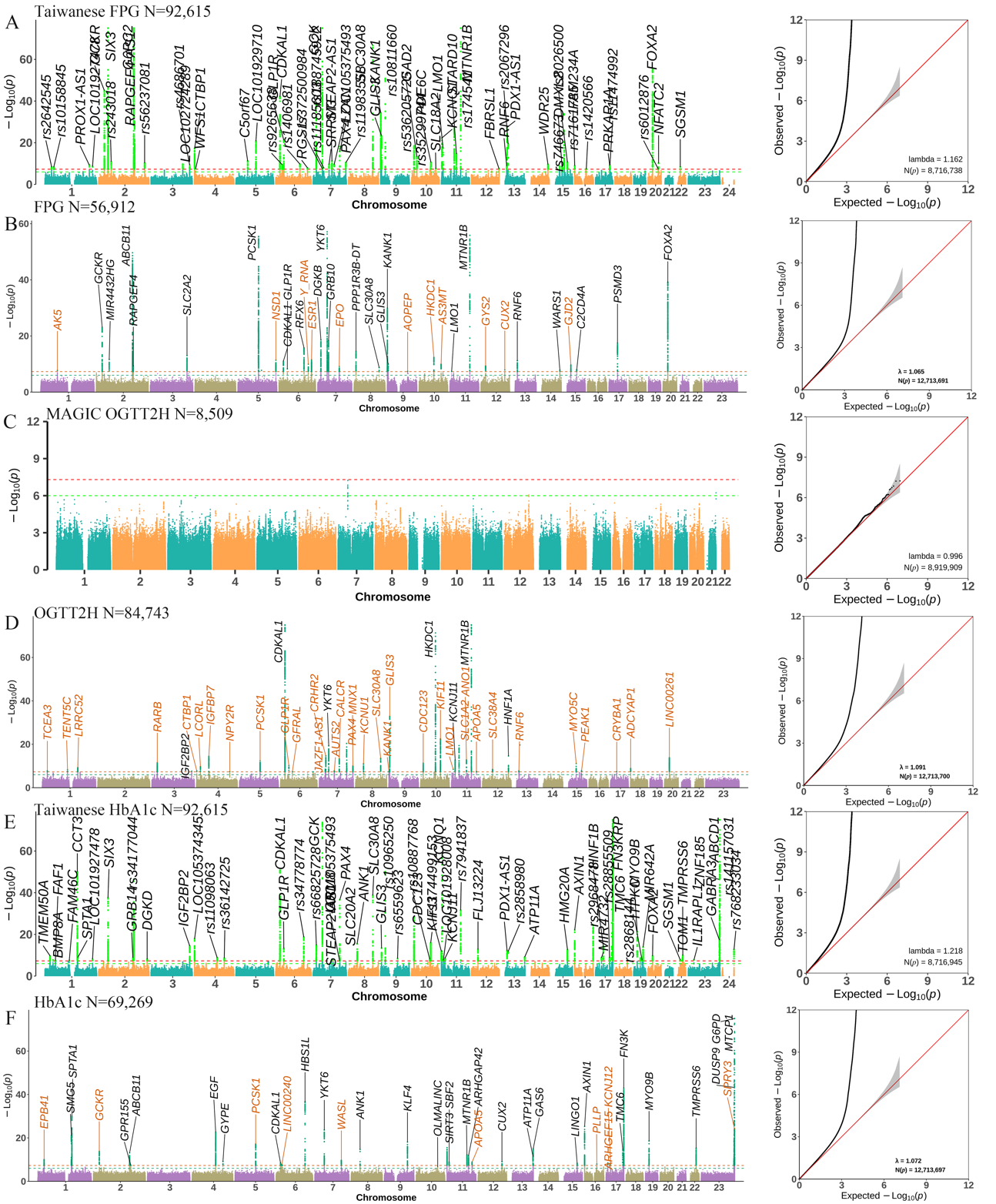

#### Figure S10. Comparison of GWAS findings from this study with the largest studies on glycemic traits in Chinese and East Asian populations.

Summary statistics for East Asian FPG and HbA1c were obtained from <https://www.ebi.ac.uk/gwas/studies/GCST90278628> and <https://www.ebi.ac.uk/gwas/studies/GCST90278632> respectively. The OGTT2H GWAS summary was downloaded from <https://magicinvestigators.org/downloads/>.

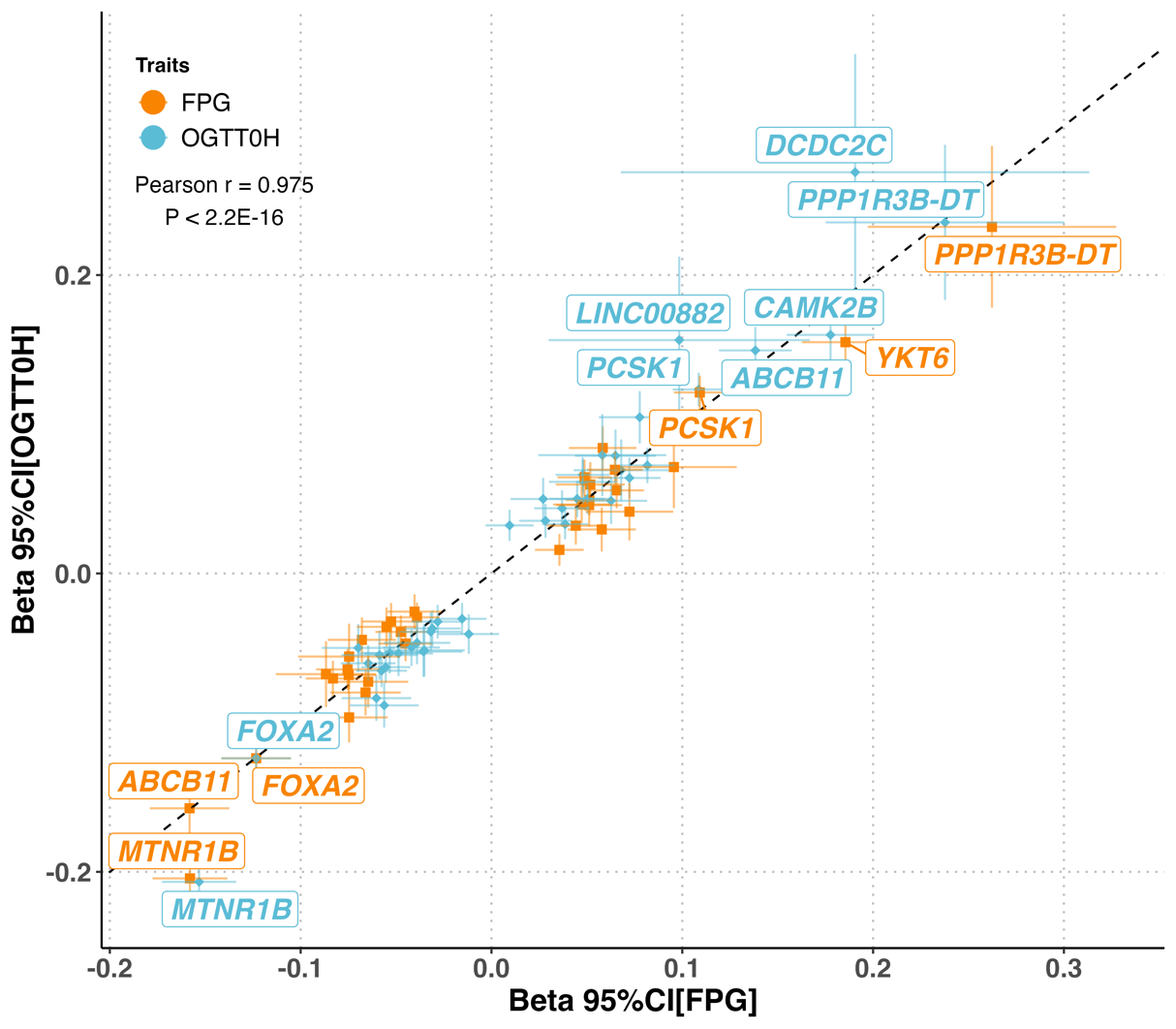

Figure S11. Comparison of lead SNP effects and genetic correlation between FPG and OGTT0H.

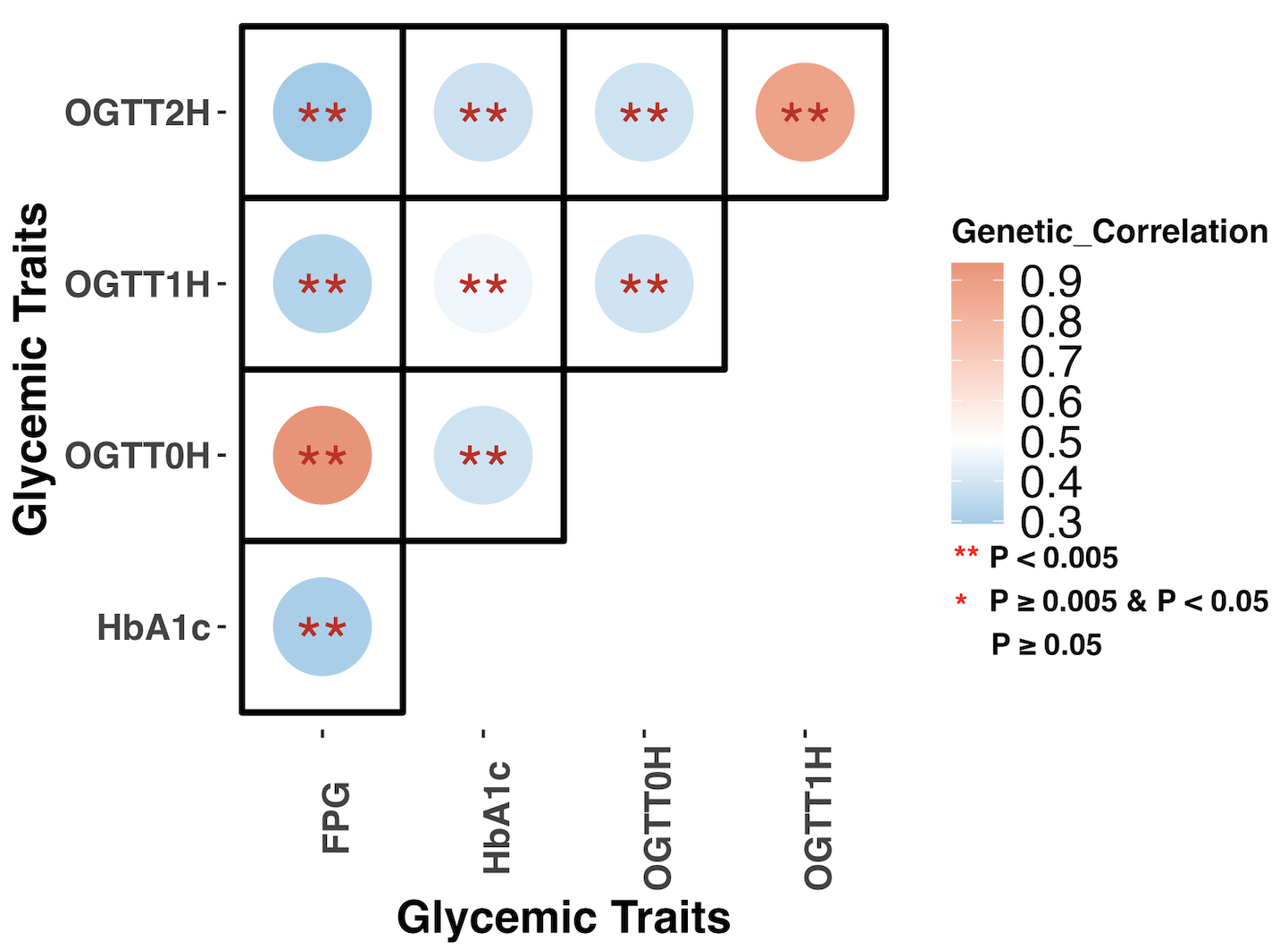

Figure S12. Genetic correlation among five glycemic traits.

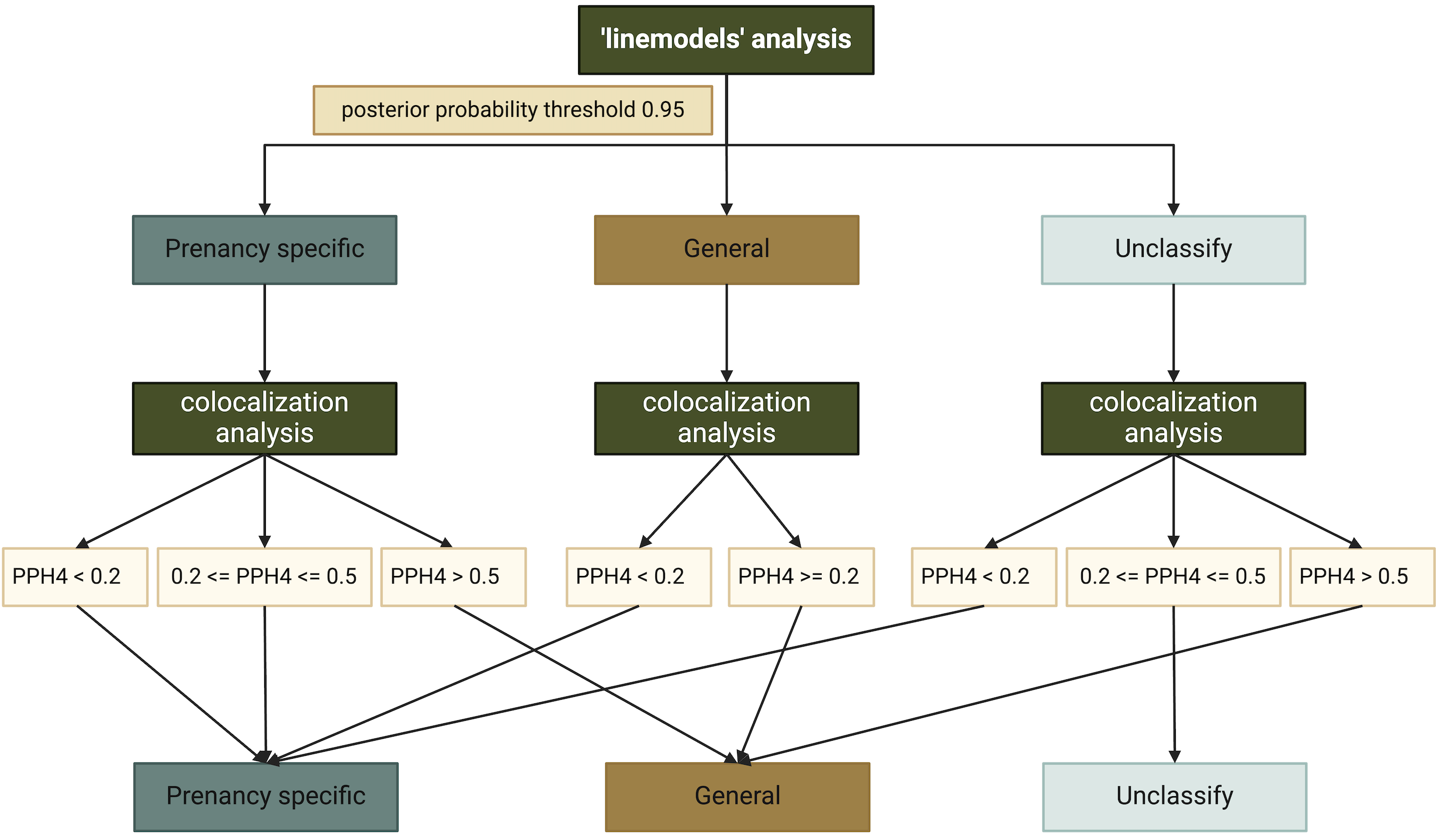

#### Figure S13. Workflow of the ‘linemodels’ approach and colocalization analysis.

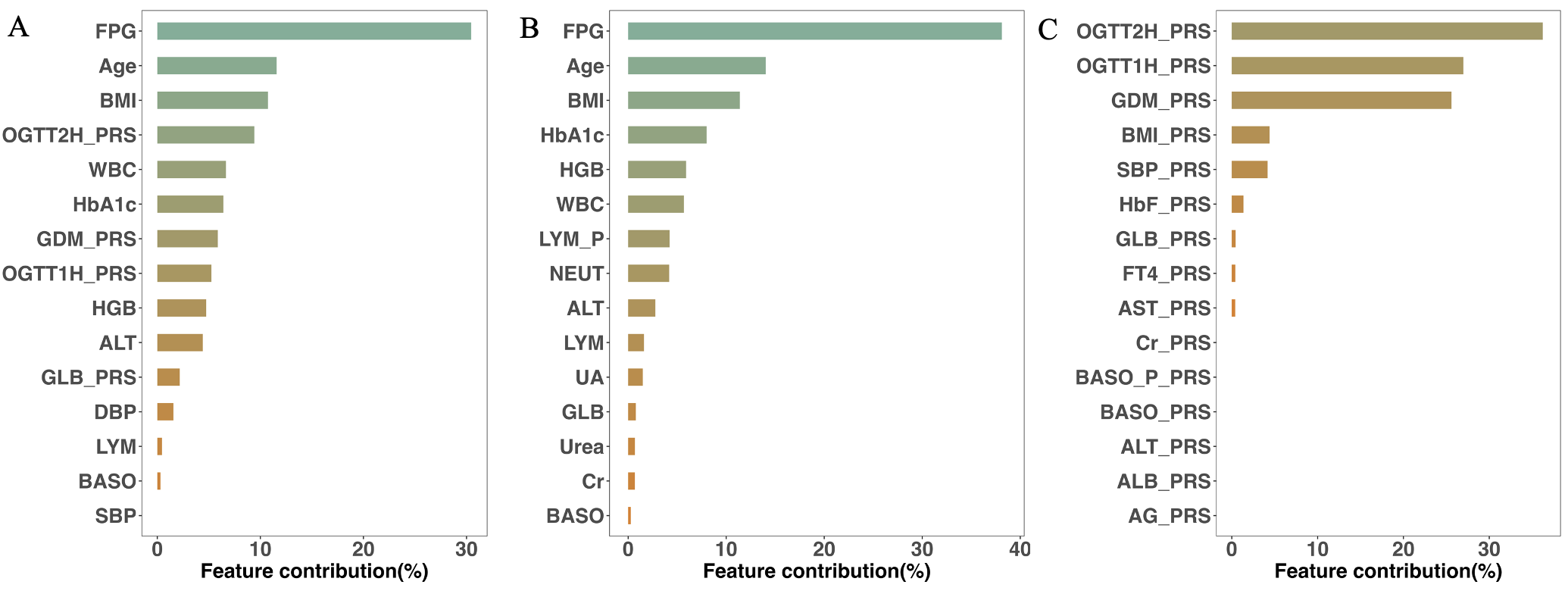

#### Figure S14. Feature contribution of the prediction model based on Shapley values.

A: Feature importance of the top 15 contributing features in the combine model. B: Feature importance of the top 15 contributing features in the non-gentic model. C: Feature importance of the top 15 contributing features in the PRS model.

Bar colors represent the influence of each feature. Full feature namea are listed in Table S1.

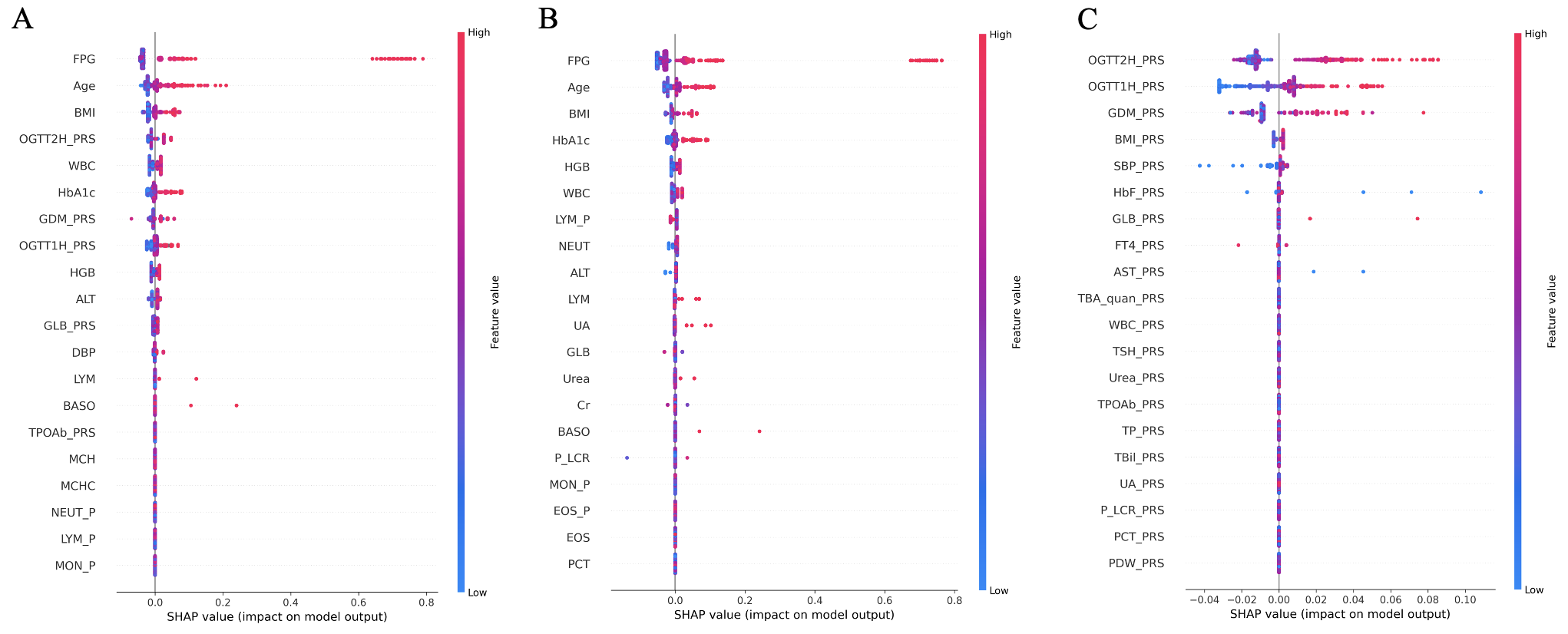

#### Figure S15. Summary plot of the prediction model based on Shapley values.

A: Summary plot of the top 20 contributing features in the combined model.

B: Summary plot of the top 20 contributing features in the non-gentic model.

C: Summary plot of the top 20 contributing features in the PRS model. Bar color trends indicate the direction of influence. Full feature names are listed in Table S1.

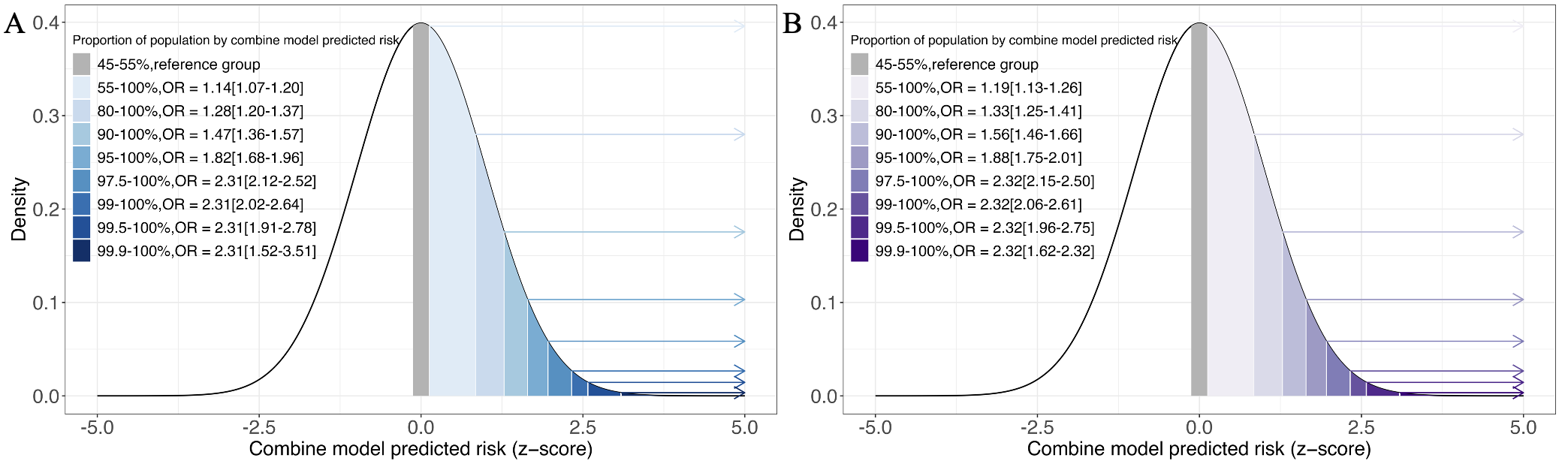

#### Figure S16. GDM risk at specific percentages in the combined model.

Panel A-B show the GDM risk at specific percentage in the combined model for validation in (A) 20% of the Baoan 20K cohort and (B) the NIPT PLUS cohort.

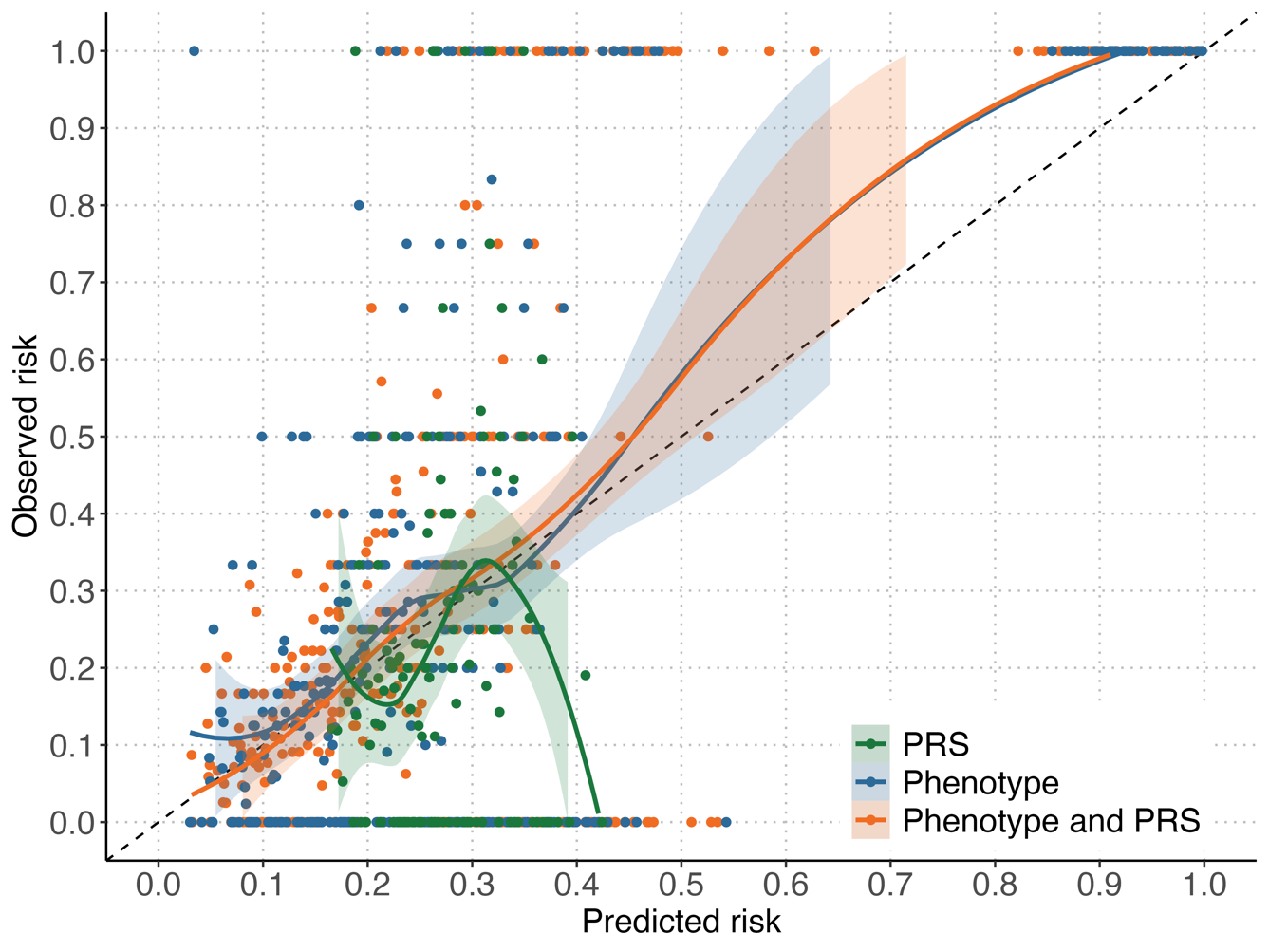

#### Figure S17. Calibration plot of the predictive model.

The calibration curves for the three predictive models demonstrate the agreement between predicted and observed risk. The dotted line represents perfect prediction, while the solid lines indicate the performance of each model.
